# Effects of non-invasive brain stimulation in dystonia: a systematic review and meta-analysis

**DOI:** 10.1101/2021.11.02.21265839

**Authors:** Jordan Morrison-Ham, Gillian M. Clark, Elizabeth G. Ellis, Andris Cerins, Juho Joutsa, Peter G. Enticott, Daniel T. Corp

**Author notes:** **Corresponding authors:** Jordan Morrison-Ham & Daniel T. Corp.

## Abstract

**Background:** Deep brain stimulation is a highly effective treatment of dystonia, but is invasive and associated with risks, such as intraoperative bleeding and infections. Previous research has used non-invasive brain stimulation (NIBS) in an attempt to alleviate symptoms of dystonia. The results of these studies, however, have been variable, leaving efficacy unclear. This study aimed to evaluate the effects of NIBS on symptoms of dystonia and determine whether methodological characteristics are associated with variability in effect size.

**Methods:** Embase and MEDLINE Complete databases were searched for articles using any type of NIBS as an intervention in dystonia patients, with changes in dystonia symptoms the primary outcome of interest.

**Results:** Meta-analysis of 26 studies demonstrated a small effect size for NIBS in reducing symptoms of dystonia (random-effects Hedges’ g = 0.21, p = .002). Differences in the type of NIBS, type of dystonia, and brain region stimulated had a significant effect on dystonia symptoms. Meta-regression revealed that 10 sessions of active stimulation, and the application of concurrent motor training programs resulted in significantly larger mean effect sizes.

**Conclusion:** NIBS has yielded small improvements to dystonic symptoms, but effect sizes depended on methodological characteristics, with more sessions of stimulation producing a larger response. Future research should further investigate the application of NIBS parallel to motor training, in addition to providing a greater quantity of sessions, to help define optimal parameters for NIBS protocols in dystonia.

**Registration:** PROSPERO 2020, CRD42020175944.

## Introduction

Dystonia is a chronic neurological disorder characterized by involuntary muscle contractions and postures^1^. Dystonia can involve any body region, and is one of the most common movement disorders, with prevalence estimates of approximately 16.43 cases per 100,000 people^2^. Dystonia can be idiopathic or secondary to other brain pathologies, such as focal brain lesions.

Invasive neuromodulation is highly effective in the treatment of dystonias^3^. Deep brain stimulation (DBS) to the globus pallidus interna (GPi) is the most widely used neuromodulation treatment for dystonia, and the subthalamic nucleus has also shown success^3, 4^. The mechanism of action for DBS in dystonia is not yet fully understood, but it is considered to modulate the function of the sensorimotor network, regions of which are often functionally abnormal in dystonia patients^5, 6^. Nevertheless, DBS is invasive and only considered in more severe cases that do not respond to botulinum toxin injections and oral pharmacotherapy^7^.

As a result, non-invasive brain stimulation (NIBS) has been suggested as a potential therapeutic treatment for dystonia symptoms, due to its ability to non-invasively modulate the functioning of abnormal neural networks^8, 9^. NIBS techniques, such as transcranial magnetic stimulation (TMS) and transcranial electrical stimulation (tES), have been effective in the treatment of other neurological and neuropsychiatric disorders, such as depression^10^, migraine^11^, and obsessive compulsive disorder^12^. Applied cortically, repetitive TMS (rTMS) and tES induce a plasticity-like response and can upregulate or downregulate neuronal activity at local and regional levels^13^.

Given that DBS in dystonia affects a large brain network, it is likely that there are multiple nodes that could be modulated via NIBS for therapeutic benefit. Several studies have demonstrated loss of inhibition, increased excitability, or abnormal plasticity in dystonia, in cortical regions associated with sensorimotor function including the somatosensory cortex (S1), primary motor cortex (M1), dorsal premotor cortex (dPM), and cerebellum^14–19^ However, NIBS to these cortical areas has returned variable results. Whilst previous research has suggested that rTMS and transcranial direct current stimulation (tDCS) provide some relief from symptoms of dystonia^20^, other studies suggest little to no effect on dystonia symptoms in comparison to sham stimulation^21, 22^. Given these conflicting results, it is not yet known whether NIBS is effective in dystonia, nor whether specific NIBS methods or brain regions may enhance therapeutic effects.

Therefore, the aim of this systematic review and meta-analysis is to pool all studies that have used NIBS in dystonia to comprehensively evaluate the effect of NIBS methods on dystonia symptoms, and to better understand which protocols may be most effective.

## Methods

### Study Selection

#### Systematic Search

A search of Embase and MEDLINE Complete was conducted during March and April 2020, using a combination of synonyms of the following terms: dystonia; transcranial magnetic stimulation (TMS); theta-burst stimulation (TBS); transcranial alternating current stimulations (tACS); transcranial direct current stimulation (tDCS); transcranial electrical stimulation (tES); transcranial random noise stimulation (tRNS); and non-invasive brain stimulation (NIBS). Exact search syntax is provided in supplementary file 1. No publication status or year limiters were applied, however only studies reported in English were considered. The reference lists of all included articles were searched for studies missed in the initial search.

#### Inclusion & Exclusion Criteria

Studies were screened using inclusion and exclusion criteria based on the PICO (participants, intervention, control, outcome) framework^23^. Studies were first selected for *qualitative review* based on the following criteria: (P) participants who had a clinical diagnosis of dystonia (any type), with a study sample size of 1 or more; (I) non-invasive brain stimulation (any type) used as an intervention intended to reduce dystonia symptom severity; (C) no comparison group or randomization necessary for qualitative review; and (O) an outcome measure that assessed changes in clinical symptoms of dystonia (e.g., Burke-Fahn-Marsden Dystonia Rating Scale).

Studies were selected for *quantitative review* (i.e., meta-analysis) based on the following criteria: (P) participants who had a clinical diagnosis of dystonia (any type), with a study sample size of at least 3; (I) as above; (C) a comparison group of dystonia controls who received sham stimulation (parallel trials), or a design where dystonia patients received both sham and real stimulation (crossover trials); (O) as above. Studies that examined dystonia participants who were actively receiving DBS were excluded, as DBS can influence the response to NIBS, even where the DBS stimulator is switched off^24, 25^.

#### Screening & Data Extraction

Literature search results were exported to EndNote (version X9) and Rayyan^26^. Two reviewers independently screened titles and abstracts obtained from the literature search against the inclusion and exclusion criteria. Full text articles were then assessed against inclusion criteria, with disagreements resolved through discussion, and where necessary by a third member of the study team (D.C.).

Following the screening and inclusion of full text articles, data were extracted from individual studies into custom Microsoft Excel spreadsheets, including participant demographics, clinical information, trial characteristics, NIBS protocols, and symptom scores. The primary outcome was changes in dystonia symptoms, post-intervention. In this review, we analysed dystonia symptoms measured by clinically validated rating scales (e.g., the Toronto Western Spasmodic Torticollis Rating Scale [TWSTRS]); subjective patient symptom scales created specifically for the empirical study; and changes in motor performance in the affected limb post-intervention. The potential influence of outcome measures on effect sizes was later analysed using meta-regression.

### Effect Size Calculations

Due to the small sample sizes of the included articles, a Hedges’ *g* effect size was calculated to correct for potential overestimation of the population standardized mean difference (SMD)^27^. For all studies, Hedges’ *g* was calculated so that positive values indicated NIBS improved dystonia symptoms, and negative values indicated NIBS worsened dystonia symptoms. Hedges’ *g* was calculated to compare the change in dystonia symptoms from baseline (either clinical or task-based) between the NIBS and sham conditions. This effect size was calculated from pre- and post-stimulation mean scores (or change from baseline scores) and standard deviations (SDs) for both NIBS and sham groups, using Comprehensive Meta-Analysis (CMA; version 3.3.070) software. In studies where the means and SDs were reported in graphs or images, Plot Digitizer (version 2.6.8; http://plotdigitizer.sourceforge.net/) software was used to extract values. If standard errors (SE) or confidence intervals (CI) were reported for mean scores, they were converted to SDs using the equations:

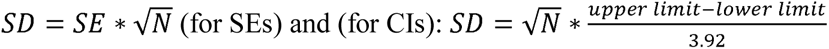

Where *N* is the total number of participants^28^. All formulas for effect size calculations are provided in Supplementary 2.

#### Pooling of Effect Sizes

For studies that used more than one outcome measure to assess symptoms of dystonia (e.g., a task-based measure along with a clinically validated rating scale)^29–34^, effect sizes and variances for each outcome were averaged within studies, to obtain one overall effect size for each study. All effect sizes were then pooled using a random effects model in CMA software. Both study level and the overall pooled effect size were considered significant if *p* < .05.

### Meta-Analysis

All meta-analysis forest plots and sensitivity analyses were conducted in Stata/SE (version 15.1). A leave-one-out sensitivity analysis was performed to detect the presence of any outliers, using the ‘metainf’ command^35^. In order to obtain an effect size estimate for each level within categorical variables, individual meta-analyses were run separating studies by NIBS type (e.g., tDCS, rTMS), brain region stimulated, type of dystonia, and outcome measures: clinically validated rating scales, unvalidated rating scales (i.e., rating scales devised for the study), and task-based outcomes (e.g., timed handwriting tests). Separate meta-analyses were conducted for each of the aforementioned variables (rather than comparing levels of the variable with a technique such as meta-regression) as there were a high number of levels per variable (e.g., high and low frequency rTMS, intermittent and continuous TBS, and tDCS for the variable NIBS type) and few study effect sizes per level, therefore insufficient statistical power to utilise a number of these variables within a meta-regression^36^.

Between-study heterogeneity in effect sizes was quantified using the *I*^2^ statistic^37^. As per Higgins^37^ the effect of heterogeneity was considered low, moderate, or high for *I*^2^ values of 25%, 50%, and 75%, respectively.

### Meta-Regression

Meta-regression analyses were conducted in Stata/SE (version 15.1) to determine the influence of mean age, gender ratio, number of active sessions of stimulation, etiology of dystonia, and concurrent motor training on NIBS outcomes. The ‘metareg’^38^ function was used for continuous variables (mean age and gender ratio), and the ‘maanova’^39^ function on the categorical variables (number of active sessions of stimulation, dystonia etiology and concurrent motor training). Prior to conducting the regression analysis, data were checked visually for normality and collinearity using histograms and scatterplots. Levels of independent variables were omitted from the regression analysis if they did not comprise at least three studies, ensuring that there were enough data for each level to provide a reliable regression estimate^36^.

### Evaluation of Bias

The methodological quality of each study was assessed using the Cochrane Collaboration’s Risk of Bias (RoB) checklists^40^. For parallel trials, the revised Cochrane Risk of Bias tool for randomized trials (RoB 2)^40^ was used, while a modified version of the RoB 2 for repeated measures designs was utilised for crossover trials. The RoB 2 checklist assesses studies on the domain’s randomization, blinding of participants and personnel, outcome measurement and assessor blinding, incomplete outcome data, and selective outcome reporting. Each domain was judged to be of low, unclear, or high risk of bias, with an overall judgement given for each study. For crossover trials, bias arising from period or carryover effects was also assessed.

The presence of publication bias across studies was assessed using funnel plots where effect sizes for each study were plotted against their standard error^41^. In the absence of publication bias, symmetrical distribution of effect sizes around the overall effect size is observed. The symmetry of the funnel plot was assessed both visually, and statistically using Egger’s test^41^.

## Results

### Study Selection

In total, 1124 records were identified across the two databases. After duplicate removal, and title and abstract screening, 121 full-text articles were assessed for eligibility. 46 studies were included for qualitative synthesis, with 26 studies (11 parallel and 15 crossover trials) meeting inclusion criteria for the meta-analysis (Figure 1).

**Figure 1.**
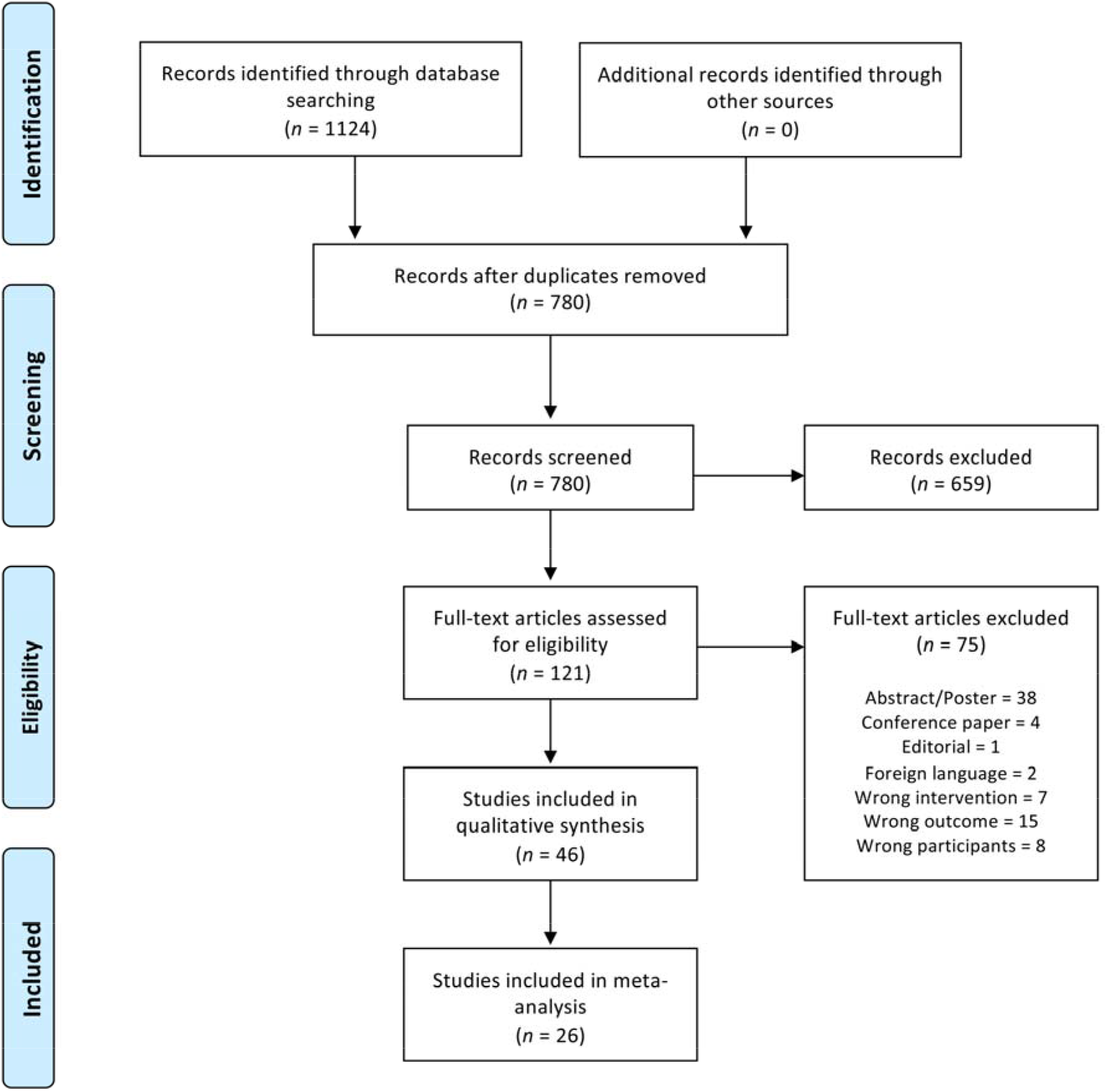
PRISMA flowchart of search method and screening process.

### Study Characteristics

A total 513 participants were included across 46 studies, with ages ranging from 7-70 years (*M* = 45.93, *SD* = 11.93). Three studies included patients with acquired dystonia, associated with Wilson’s disease^43^ or cerebral palsy^44, 45^. Thirteen studies included participants who were not on oral medications (e.g., benzodiazepines), and 18 studies examined participants who had their last botulinum toxin injection more than 4 weeks prior to stimulation. Low frequency rTMS (1Hz or 0.2Hz; 21 studies) was the most utilised form of NIBS, followed by tDCS (anodal or cathodal; 17 studies). A single study applied tACS^50^. Dystonia-specific motor training or biofeedback was employed concurrently with NIBS in seven studies^29, 30, 32, 33, 46–48^. All study designs, participant demographics and characteristics are provided in Table 1.

**Table 1.** Included articles’ participant demographics and characteristics, study designs and outcomes.

| Study | Design | Participants | Dystonia Cause | Intervention | Stimulation Target | Outcome Measure | Symptom change after NIBS |
| --- | --- | --- | --- | --- | --- | --- | --- |
| *Allam et al. (2007)*<sup>49</sup> | Case study | 1 CD/WC patient (37 years) | Idiopathic | 5 sessions rTMS (1Hz, 1200 stimuli @ 90% RMT – 20 mins) | Left dPM | BFMDRS | CD ↓ for 4 months<br>WC no change |
| *Angelakis et al. (2013)*<sup>50</sup> | Sham-controlled case study | 1 CD patient (54 years) | Idiopathic | a. 5 real, 5 sham sessions tDCS (1.5mA – 15 mins) | a. Cathode C4, anode P3 | TWSTRS, patient rating scale | ↓ |
|  |  |  |  | b. 7 sessions tACS (15Hz, 5Hz, 15Hz – 6/3/6 mins) | b. SMC |  | TWSTRS ↓ 29.5pts |
| *Benninger et al. (2011)*<sup>21</sup> | RCT | 12 WC patients ($M = 57.1$) | Idiopathic | 3 sessions real or sham cathodal tDCS (2mA – 20 mins) | Contralateral M1 | ADDS, WCRS, handwriting kinematics | ↑ |
| *Betti et al. (2018)*<sup>51</sup> | Case study | 1 MD patient (55 years) and 1 HC (50 years) | Idiopathic | 5 sessions rTMS (1Hz, 1800 stimuli @ 90% RMT – 30 mins) | Left M1 | Motor tasks | ↓ |
| *Bhanpuri et al. (2015)*<sup>44</sup> | Crossover RCT | a. 7 HD patients ($M = 15.7$) | a. 1 DYT1, 6 acquired (4 cerebral palsy, 1 vitamin E deficient, 1 TBI) | a. 5 real, 5 sham sessions cathodal tDCS (2mA – 9 mins) | a. Contralateral M1 | BADs, EMG tracking task | ↓ |
| | | b. 6 HD patients ($M = 14.8$) | b. 6 acquired (1 TBI, 5 cerebral palsy) | b. 5 real, 5 sham sessions anodal tDCS (2mA – 9 mins) | b. Contralateral M1 | | ↑ |
| *Bologna et al. (2016)*<sup>52</sup> | Crossover RCT | 13 FHD ($M = 48.5$), 13 CD ($M = 46.7$) and 13 HC ($M = 49.9$) | Idiopathic | 1 real, 1 sham session cTBS (600 stimuli – 40 secs) | Cerebellum (ipsilateral to affected hand in FHD) | TWSTRS, WCRS, cortical excitability | No change |
| *Borich et al. (2009)*<sup>53</sup> | Crossover RCT | 6 FHD patients ($M = 46.5$) and 9 HC ($M = 33$) | Idiopathic | 5 sessions rTMS (1Hz, 900 stimuli @ 90% RMT), 3 patients received sham before crossing over to the intervention | Contralateral dPM | Handwriting kinematics, cortical excitability | Handwriting (pen velocity) ↓* |

NIBS in Dystonia: A Meta-Analysis
| Study | Design | Participants | Dystonia Cause | Intervention | Stimulation Target | Outcome Measure | Symptom change after NIBS |
| --- | --- | --- | --- | --- | --- | --- | --- |
| *Bradnam et al. (2014)*<sup>54</sup> | Case study | 1 CD patient (47 years) | Idiopathic | 20 sessions anodal tDCS (2mA – 2 x 15 min blocks, separated by 5 mins) | Right/left cerebellum (5 sessions each), right M1/left cerebellum (10 sessions) | TWSTRS, CDQ-24, CDIP-58 | TWSTRS, CDQ-24, CDIP-58 ↓ 18+ pts |
| Bradnam et al. (2015)<sup>55</sup> | Crossover RCT | 8 FHD patients ($M = 59$) and 8 HC ($M = 61.3$) | Idiopathic | 1 session each anodal, cathodal, and sham tDCS (2mA – 20mins) | Cerebellum | Handwriting kinematics, cortical excitability | Anodal tDCS handwriting ↓* |
| Bradnam et al. (2016)<sup>8</sup> | RCT | 16 CD patients ($M = 51.9$) | Idiopathic | 10 sessions real or sham iTBS (600 stimuli each hemisphere – 8 mins) | Bilateral cerebellum | TWSTRS, CDQ-24, hand dexterity, cortical excitability | TWSTRS, CDQ-24, hand dexterity ↓* |
| Buttkus et al. (2010a)<sup>56</sup> | Crossover RCT | 10 MD patients ($M = 48.8$) | Idiopathic | 1 real, 1 sham session cathodal tDCS (2mA – 20 mins) | Left M1 | ADDS, FAM, assessment of fine motor control | No change |
| *Buttkus et al. (2010b)*<sup>47</sup> | Case study | 1 MD patient (43 years) | Idiopathic | 5 cathodal, 5 anodal, 5 sham sessions tDCS + training (2mA – 20 mins) | Left M1 | Assessment of fine motor control | Anodal, cathodal tDCS ↓* |
| Buttkus et al. (2011)<sup>29</sup> | Crossover RCT | 9 MD patients ($M = 44$) | Idiopathic | 1 cathodal, 1 anodal, 1 sham session tDCS + training (2mA – 20mins) | Left M1 | Assessment of fine motor control | No change |
| *Conte et al. (2014)*<sup>57</sup> | Crossover RCT | 12 WC patients ($M = 43$) and 12 HC ($M = 42$) | Idiopathic | 1 iTBS, 1 cTBS, 1 sham session (iTBS = 600 stimuli @ 80% AMT, cTBS = 600 stimuli @ 80% AMT – 40 secs) | Contralateral S1 | Writing task, STDT | No change |
| Furuya et al. (2014)<sup>30</sup> | Crossover RCT | 10 MD patients ($M = 39.6$) and 10 HC ($M = 27.9$) | Idiopathic | 1 cathodal, 1 anodal, 1 anodal over unaffected hemisphere, 1 sham session (all + training), and 1 cathodal session (no training), 5 sessions total (2mA – 24 mins) | Contralateral M1, except for 1 anodal session over ipsilateral M1 | Assessment of fine motor control | ↓ for cathodal (+training) only* |
| Havrankova et al. (2010)<sup>58</sup> | Crossover RCT | 11 WC patients ($M = 40.3$) | Idiopathic | 5 real, 5 sham sessions rTMS (1Hz, 1800 stimuli @ 90% AMT – 30 mins) | Contralateral S1 | BFMDRS, handwriting, 2-minute writing test | Handwriting ↓* |

NIBS in Dystonia: A Meta-Analysis
| Study | Design | Participants | Dystonia Cause | Intervention | Stimulation Target | Outcome Measure | Symptom change after NIBS |
| --- | --- | --- | --- | --- | --- | --- | --- |
| Huang *et al.* (2010)<sup>59</sup> | RCT | 7 patients ($M = 44.9$) and 9 HC ($M = 42.7$) | Idiopathic | 1 session cTBS (600 stimuli @ 80% AMT – 40 secs) | Contralateral dPM | Handwriting, cortical excitability | Handwriting ↓* |
| Huang *et al.* (2012)<sup>60</sup> | RCT | 18 WC patients ($M = 41.4$) | Idiopathic | 5 sessions real or sham cTBS (600 stimuli @ 80% AMT – 40 secs) | Contralateral dPM | Handwriting, cortical excitability | No change |
| Kimberley *et al.* (2013)<sup>61</sup> | RCT | 17 FHD patients ($M = 46.5$) | Idiopathic | 5 sessions real or sham rTMS (1Hz, 1800 stimuli @ 90% RMT – 30 mins) | Contralateral dPM | Handwriting kinematics, cortical excitability | Handwriting (pen pressure) ↓* |
| Kimberley *et al.* (2015a)<sup>48</sup> | Crossover RCT | 9 FHD patients ($M = 46$) | Idiopathic | 5 sessions rTMS + training, 5 sessions rTMS only (1Hz, 1200 stimuli @ 80% RMT – 20 mins) | Contralateral dPM | ADDS, GROC, WCRS, handwriting kinematics, cortical excitability | ADDS ↓ for both conditions* |
| Kimberley *et al.* (2015b)<sup>62</sup> | Case study | 2 FHD patients ($M = 43.5$) | Idiopathic | 6 sessions rTMS (1Hz, 1200 stimuli @ 90% RMT – 20 mins) | Right dPM | Handwriting kinematics, cortical excitability, patient rating scale | Handwriting ↓* |
| Koch *et al.* (2014)<sup>9</sup> | RCT | 20 CD patients ($M = 54$) and 10 HC<sup>†</sup> ($M = 51.2$) | Idiopathic | 10 sessions real or sham cTBS (600 stimuli per hemisphere @ 80% AMT – 4 mins) | Bilateral cerebellum | TWSTRS, BFMDRS, cortical excitability | TWSTRS ↓* |
| Kranz *et al.* (2009)<sup>63</sup> | Randomised crossover study | 7 BEB patients ($M = 62.6$) | Idiopathic | a. 4 sessions rTMS (0.2Hz, 180 stimuli @ 90% RMT – 15 mins) | a. M1, dPM, SMA, ACC | Physician/patient rating scales, BRR | Rating scales ↓ for all targets* |
|  |  |  |  | b. 4 sessions cTBS (600 stimuli @ 80% AMT – 40 secs) | b. M1, dPM, SMA, ACC |  | No change |
|  |  |  |  | c. 2 sessions tDCS (1mA – 20 mins) | c. M1/dPM, SMA/ACC |  | No change |
| Kranz *et al.* (2010)<sup>64</sup> | Crossover RCT | 12 BEB patients ($M = 61.4$) | Idiopathic | 2 rTMS (H-coil and C-coil), 1 sham session (0.2Hz, 180 stimuli @ 100% AMT – 15 mins) | ACC | Physician/patient rating scales, BRR | Rating scales, BRR ↓ for both coils* |

NIBS in Dystonia: A Meta-Analysis
| Study | Design | Participants | Dystonia Cause | Intervention | Stimulation Target | Outcome Measure | Symptom change after NIBS |
| --- | --- | --- | --- | --- | --- | --- | --- |
| *Lefaucher et al. (2004)*<sup>65</sup> | Case series | 3 CD/lower limb dystonia patients ($M = 38.3$) | Acquired (1 viral encephalitis, 1 neonatal TBI, 1 disulfiram intoxication) | 5 sessions rTMS (1Hz, 1200 stimuli @ 90% RMT – 20 mins) | Left dPM | BFMDRS, spasm number/intensity | Spasm number/intensity ↓ |
| Linssen et al. (2015)<sup>66</sup> | Crossover RCT | 10 WC patients and 10 HC<sup>†</sup> | Not provided | 1 real, 1 sham cTBS session (80% AMT) | Ipsilateral cerebellum | Writing tasks | No change |
| Lozeron et al. (2017)<sup>43</sup> | Crossover RCT | 13 FHD patients ($M = 42.3$) | Inherited (Wilson's Disease) | 1 real, 1 sham session rTMS (1Hz, 1200 stimuli @ 80% RMT – 20 mins) | Contralateral S1 | WCRS, FAR, UWDRS | No change |
| *Marceglia et al. (2017)*<sup>67</sup> | Sham-controlled crossover study | 2 MD patients ($M = 41$) | Idiopathic | 5 cathodal, 5 anodal, 5 sham sessions tDCS (2mA – 20mins) | Bilateral M1/dPM | SSS, FSS, TC, writing tasks | Cathodal tDCS ↓ |
| Murase et al. (2005)<sup>68</sup> | Crossover RCT | 9 WC patients ($M = 38$) and 7 HC<sup>†</sup> ($M = 36$) | Idiopathic | 3 real, 1 sham session rTMS (0.2Hz, 180 stimuli @ 85% RMT/AMT for SMA) | Contralateral M1, dPM, SMA (1 session each) | Handwriting, cortical excitability | ↓ for dPM target* |
| *Naro et al. (2019)*<sup>69</sup> | Case study | 1 WC patient (25 years) | Idiopathic | 75 sessions rTMS (1Hz, 1200 stimuli @ 90% AMT) | Left dPM | WCRS, 1 minute writing test | ↓ for 12 months |
| Odorfer et al. (2019)<sup>70</sup> | RCT | 8 CD patients and 8 HC | Idiopathic | 2 real, 1 sham session cTBS (600 stimuli @ 80% AMT – 40 secs) | Left dPM, bilateral cerebellum (1 session each) | TWSTRS, cortical excitability | No change |
| *Okada et al. (2018)*<sup>49</sup> | Case study | 1 WC patient (47 years) | Idiopathic | 6 sessions tDCS (2mA – 30 mins) +EMG biofeedback | Bilateral M1 | WCRS | ↓ |
| Pirio Richardson et al. (2015)<sup>31</sup> | Crossover RCT | 9 CD patients ($M = 53$) | Idiopathic | 4 real, 1 sham session rTMS (0.2Hz, 180 stimuli @ 85% RMT – 15 mins) | Left ACC, dPM, M1, SMA (1 session each) | TWSTRS, cortical excitability | ↓ dPM, SMA, M1 targets |
| Rossett-Llobet et al. (2014)<sup>32</sup> | RCT | 4 FHD patients ($M = 35.5$) and 30 FHD controls ($M = 34.4$) | Idiopathic | 10 sessions cathodal tDCS +training/10 sessions training only, or 20 sessions cathodal tDCS +training (2mA – 20 mins) | Contralateral M1 | Motor tasks, patient rating scale | 20 sessions cathodal tDCS +training ↓* |

NIBS in Dystonia: A Meta-Analysis
| Study | Design | Participants | Dystonia Cause | Intervention | Stimulation Target | Outcome Measure | Symptom change after NIBS |
| --- | --- | --- | --- | --- | --- | --- | --- |
| Rossett-Llobet et al. (2015)<sup>33</sup> | RCT | 26 MD patients ($M = 35$) | Idiopathic | 10 session real or sham cathodal tDCS + training (2mA – 20 mins) | Left M1 | Dystonia severity rating, therapist rating | ↓* |
| Sadnicka et al. (2014)<sup>22</sup> | Crossover RCT | 10 WC patients ($M = 52.8$) | Idiopathic | 1 real, 1 sham session anodal tDCS (2mA – 15 mins) | Cerebellum | WCRS, cortical excitability | No change |
| Salatino et al. (2019)<sup>71</sup> | Sham-controlled case study | 1 FHD patient (41 years) and 1 HC<sup>†</sup> (32 years) | Idiopathic | a. 1 real, 1 sham rTMS sessions (1Hz, 900 stimuli @ 90% RMT) | Left dPM | Handwriting, therapist rating | Handwriting ↓ |
|  |  |  |  | b. 6 sessions rTMS (1Hz, 900 stimuli @ 90% RMT) |  |  | Handwriting ↓ |
| Sharma et al. (2019)<sup>72</sup> | Case study | 1 lower limb dystonia patient (65 years) | Idiopathic | 10 sessions rTMS (1Hz, 2600 stimuli 90% AMT – 44 mins) | Right M1 | PHQ-9, CGI | No change |
| Shin et al. (2019)<sup>73</sup> | Case study | 1 lower limb dystonia patient (57 years) | Acquired (cerebellar lesion) | 5 sessions rTMS (1Hz, 600 stimuli @ 90% RMT) | Right cerebellum | BFMDRS | ↓ |
| Siebner et al. (1999)<sup>34</sup> | Crossover RCT | 16 WC patients ($M = 47$) and 11 HC ($M = 40$) | Idiopathic | 1 session rTMS (1Hz, 1800 stimuli @ 90% RMT – 30 mins), 10 WC patients received sham before crossing over to the intervention | Left M1 | Handwriting kinematics, cortical excitability | Handwriting pressure ↓* |
| Siebner et al. (2003)<sup>74</sup> | Crossover RCT | 7 FHD patients ($M = 48$) and 7 HC ($M = 48$) | Idiopathic | 1 real, 1 sham session rTMS (1Hz, 1800 stimuli @ 90% RMT – 2x15 min trains) | Left dPM | WCRS, physician rating scale, handwriting | No change |
| Trebbosson et al. (2017)<sup>75</sup> | Case study | 1 BEB patient (70 years) | Not provided | 10 sessions tDCS (2mA – 30 mins) | Left dPFC | Eye blink rate, depressive symptom report | ↓ for 10 days |
| Veugen et al. (2013)<sup>76</sup> | RCT | 15 WC patients ($M = 58$) and 10 HC ($M = 58$) | Idiopathic | 1 session cTBS (600 stimuli @ 70% RMT) | Contralateral dPM | Writing tasks, cortical excitability | Handwriting (spiral maze) ↓* |

NIBS in Dystonia: A Meta-Analysis
| Study | Design | Participants | Dystonia Cause | Intervention | Stimulation Target | Outcome Measure | Symptom change after NIBS |
| --- | --- | --- | --- | --- | --- | --- | --- |
| *Vucurovic et al. (2017)*<sup>77</sup> | Case study | 1 left multifocal dystonia patient (41 years) | Acquired (Schizencephaly-related left hemiparesis with spastic dystonia) | 3 sessions rTMS (1Hz, 1200 stimuli @ 100% RMT) | Left M1 | UDRS | ↓ for 1 month |
| Wagle-Shukla et al. (2018)<sup>78</sup> | RCT | 12 BEB patients ($M = 69.1$) | Not provided | 10 sessions real or sham rTMS (0.2Hz, 180 stimuli @ 100% AMT – 15 mins) | ACC | JRS, CDQ-24, BRR | JRS, CDQ-24 ↓ for 2 weeks* |
| Young et al. (2014)<sup>45</sup> | Crossover RCT | 14 HD patients ($M = 12.6$) | 2 idiopathic, 12 acquired (11 cerebral palsy, 1 TBI) | 1 real, 1 sham session cathodal tDCS (1mA – 2x9 mins, with 20 min break in between) | Contralateral M1 | BADS, EMG tracking task | EMG task (overflow) ↓* |

*Note.* Where boxes are left blank, the information was not provided. Where authors are italicised, studies were included in qualitative analysis only. WC = writer's cramp; rTMS = repetitive transcranial magnetic stimulation; RMT = resting motor threshold; BFMDRS = Burke-Fahn-Marsden Dystonia Rating Scale; SMC = sensorimotor cortex; TWSTRS = Toronto Western Spasmodic Torticollis Rating Scale; RCT = randomised controlled trail; ADDS = Arm Dystonia Disability Scale; WCRS = Writer's Cramp Rating Scale; MD = Musician's dystonia; HC = healthy control; HD = hand dystonia; TBI = traumatic brain injury; BADS = Barry-Albright Dystonia Rating Scale; EMG = electromyogram; cTBS = continuous theta-burst stimulation; CDQ-24 = Craniocervical Dystonia Questionnaire; CDIP-58 = Cervical Dystonia Impact Profile; iTBS = intermittent theta-burst stimulation; FAM = frequency of abnormal movements scale; AMT = active motor threshold; STDT = sensory-temporal discrimination task; GROG = Global Rating of Change; BEB = Benign Essential Blepharospasm; SMA = supplementary motor area; BRR = blink reflex recovery; FAR = flow, accuracy and rhythmicity evaluation; UWDRS = Unified Wilson's Disease Rating Scale; SSS = Symptom Severity Scale; FSS = Functional Status Scale; TC = Tubiana and Champagne scale; PHQ-9 = Patient Health Questionnaire; CGI = Clinical Global Impression; dPFC = dorsolateral prefrontal cortex; UDRS = Unified Dystonia Rating Scale; JRS = Jankovic Rating Scale.
\* denotes statistically significant results in comparison to baseline or sham condition ( $p < .05$ ). † refers to participants used for electrophysiological data comparison only. ↓ refers to a decrease in symptoms after NIBS on outcome measures. ↑ refers to an increase in symptoms after NIBS on outcome measure.

### Qualitative Analysis

20 studies met criteria for qualitative analysis only, encompassing 73 participants with dystonia and 32 healthy control subjects (Table 1 – see italicized author studies). Overall, 19 of the 20 studies reported some reduction in dystonia symptoms after the application of NIBS, however many did not report whether this was statistically significant. Two studies^72, 73^ applied rTMS to patients with lower limb dystonia, and one study applied rTMS in a patient with left side multifocal dystonia, which affected the upper and lower limbs^77^.The average number of stimulation sessions was 10 (*SD* = 15.1), with a maximum of 75 sessions^69^.

### Meta-Analysis

Meta-analysis was performed on 26 studies, totaling 352 participants with dystonia (hand dystonias, inclusive of task-specific FHD, musician’s dystonia and writer’s cramp, 19 studies; CD, 5 studies; blepharospasm, 2 studies). Included studies were either parallel (*n* = 11; where participants were randomly assigned to sham or intervention groups) or crossover (*n* = 15; where participants completed both sham and intervention conditions) group designs. One crossover group study^74^ only provided post-stimulation data, and thus was treated as a parallel group design. Participant mean age was 44.6 years (*SD* = 13.7). The mean number of sessions of stimulation was 4.61 (*SD* = 3.57), inclusive of sham stimulation sessions in crossover trials. Of the 26 studies, 15 showed a reduction in dystonia symptoms after the application of NIBS.

Prior to conducting the meta-analysis, a leave-one-out sensitivity analysis was performed, demonstrating the presence of an outlier^64^ (Supplementary 3). This study was therefore removed from all subsequent analyses. Nevertheless, meta-analysis conducted with this study^64^ included was still significant (Supplementary 4).

Overall meta-analysis demonstrated a small effect size favoring active stimulation over sham stimulation for a reduction in dystonia symptoms, random-effects Hedges’ *g* = 0.21, 95% CI [0.08, 0.35], *p* = .002 (Figure 2). Between-study heterogeneity was significant (*I^2^* = 45.04%, *p* = .012), therefore meta-regressions were conducted to find moderators of the effect.

**Figure 2.**
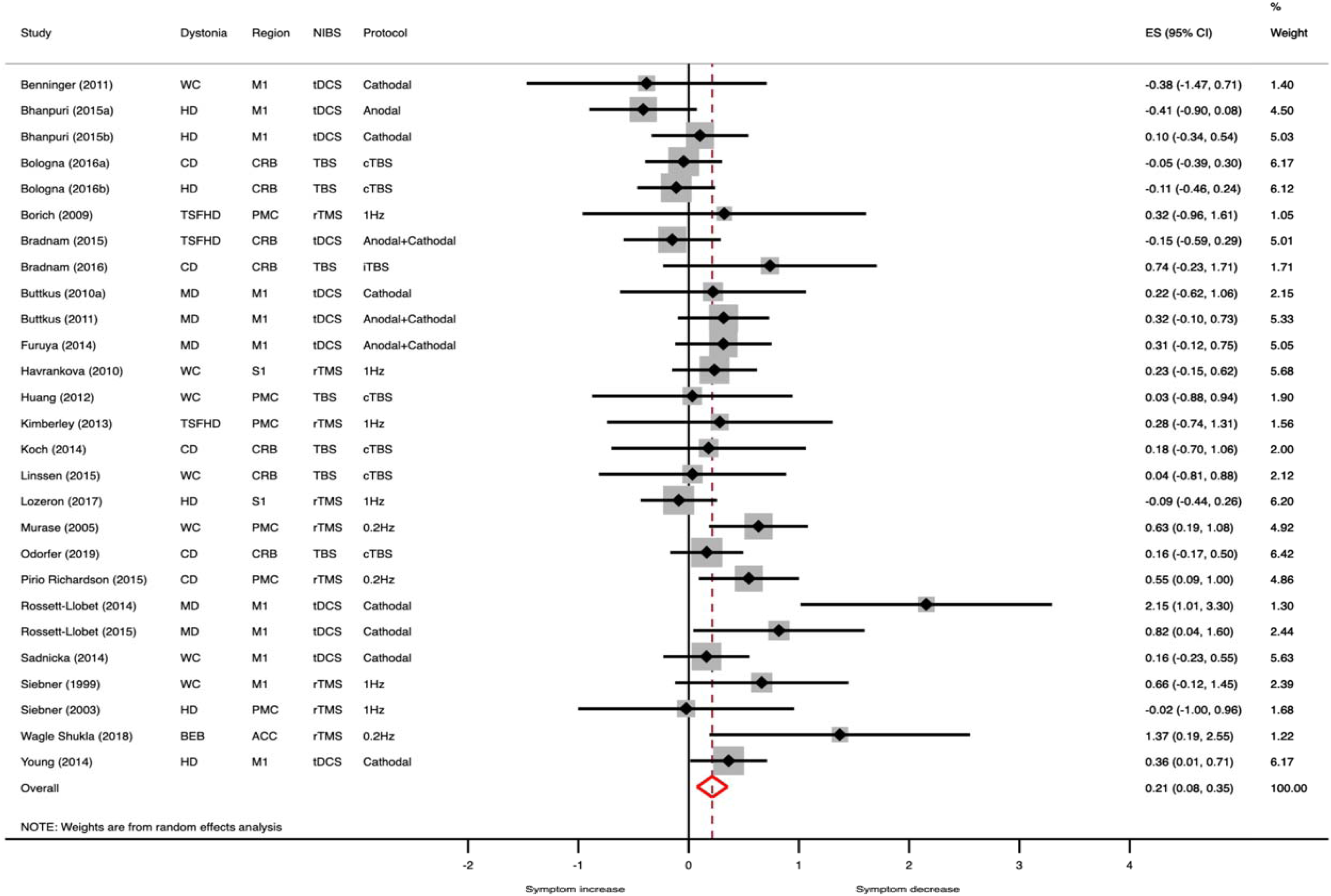
Forest plot of the random effects meta-analysis, demonstrating a small, significant effect for NIBS in decreasing dystonia symptoms. Where protocol states “Anodal+Cathodal”, participants received both anodal and cathodal tDCS. Separate effect sizes were calculated for each protocol and then combined into one overall study effect size.

Meta-analyses were then run separating studies by selected variables (Table 2). These analyses demonstrate significance for rTMS overall (*p* = .002), 0.2Hz rTMS (*p* < .001), cathodal tDCS (*p* = .04), brain regions ACC (*p* = .02), M1 (*p* = .03) and dPM (*p* = .001), and blepharospasm (*p* = .02), task-specific FHD (*p* = .002), musician’s dystonia (p = .01), and writer’s cramp (*p* = 0.007). All forest plots are available in the supplementary materials.

**Table 2.** Effect sizes for separate meta-analyses on categorical variables.

| Variable | *n* | Hedges' *g* | 95% CI | *I*<sup>2</sup> |
| --- | --- | --- | --- | --- |
| **NIBS type** |  |  |  |  |
| rTMS | 9 | 0.36* | [0.1, -0.61] | 36.4% |
| 1Hz | 6 | 0.12 | [-0.12, 0.35] | 0% |
| 0.2Hz | 3 | 0.64* | [0.36, 0.95] | 0% |
| TBS | 7 | 0.04 | [-0.14, 0.23] | 0% |
| *iTBS* | 1 | 0.74 | [-0.23, 1.71] | - |
| *cTBS* | 6 | 0.02 | [-0.17, 0.24] | 0% |
| tDCS | 11 | 0.22 | [-0.03, 0.47] | 59.8%* |
| *Cathodal* | 7 | 0.38* | [0.02, 0.74] | 59.4%* |
| *Anodal* | 1 | -0.41 | [-0.90, 0.08] | - |
| *Anodal+Cathodal* | 3 | 0.17 | [-0.13, 0.47] | 31.5%* |
| **Brain region** |  |  |  |  |
| ACC | 1 | 1.37* | [0.19, 2.55] | - |
| CRB | 7 | 0.02 | [-0.16, 0.19] | 0% |
| M1 | 11 | 0.29* | [0.04, 0.55] | 57.3%* |
| dPM | 6 | 0.46* | [0.19, 0.73] | 0% |
| S1 | 2 | 0.06 | [-0.25, 0.38] | 32.8% |
| **Dystonia type** |  |  |  |  |
| BEB | 1 | 1.37* | [0.19, 2.55] | - |
| CD | 5 | 0.22 | [-0.03, 0.47] | 25.2% |
| HD | 6 | -0.01 | [-0.23, 0.21] | 35.5% |
| Task-specific FHD | 15 | 0.32* | [0.11, 0.52] | 35.3% |
| *MD* | 5 | 0.6* | [0.13, 1.07] | 61.9%* |
| *WC* | 7 | 0.28* | [0.08, 0.49] | 0% |
| **Outcome type** |  |  |  |  |
| Validated scale | 13 | 0.17 | [-0.05, 0.38] | 54.8%* |
| Unvalidated scale | 2 | 1.06 | [-1.05, 3.17] | 87.3%* |
| Task-based measure | 20 | 0.13 | [-0.01, 0.26] | 29.5% |
| Overall | 27 | 0.21* | [0.08, 0.35] | 42.3%* |

*Note.* *I*<sup>2</sup> statistics were not calculated for several variables due to there only being one study. Three studies that assessed task-specific FHD could not be separated further into musician's dystonia and writer's cramp, as the studies included both participants. CRB = cerebellum; BEB = blepharospasm; HD = hand dystonia; MD = musician's dystonia; WC = writer's cramp. \*denotes significance at the *p* < .05 level

### Meta-Regression

Meta-regression conducted on the number of active sessions of stimulation demonstrated a significant difference between the three groups, *Q*(2) = 10.97, *p* = .004. Pairwise comparisons revealed 10 sessions of active stimulation resulted in significantly larger mean effect sizes for NIBS reducing dystonia symptoms (one session *g* = 0.2, *p* = .01, five sessions *g* = 0.04, *p* = .77, 10 sessions *g* = 0.92, *p* < .001; Figure 3). Two and three sessions of stimulation were removed as they did not meet the number of studies to be included in the analysis (*n* ≥ 3).

**Figure 3.**
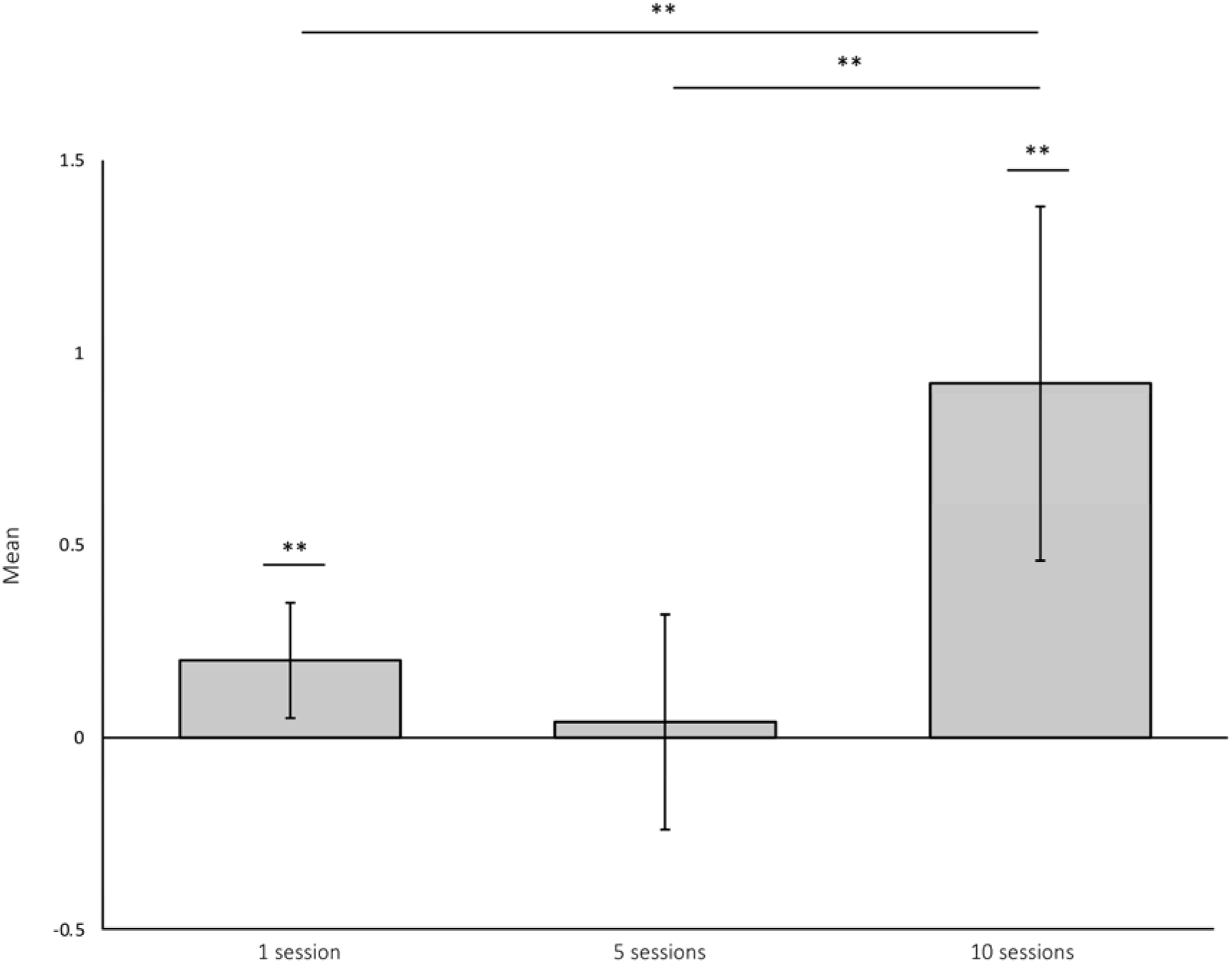
Meta-regression and pairwise comparisons conducted on the number of active sessions of stimulation. Significant differences were found between 1 and 10 sessions, and 5 and 10 sessions of stimulation. **denotes significance at the *p* < .05 level.

There were no significant differences between idiopathic and acquired dystonia study effect sizes (*Q*(1) = 2.12, *p* = .13), although idiopathic dystonia studies displayed a significant mean effect (idiopathic *g* = 0.26, *p* < .001), whereas acquired studies did not (*g* = 0.02, *p* = 0.89; Figure 4). The lack of significant difference in pairwise comparisons is likely due to the small number of acquired dystonia studies.

**Figure 4.**
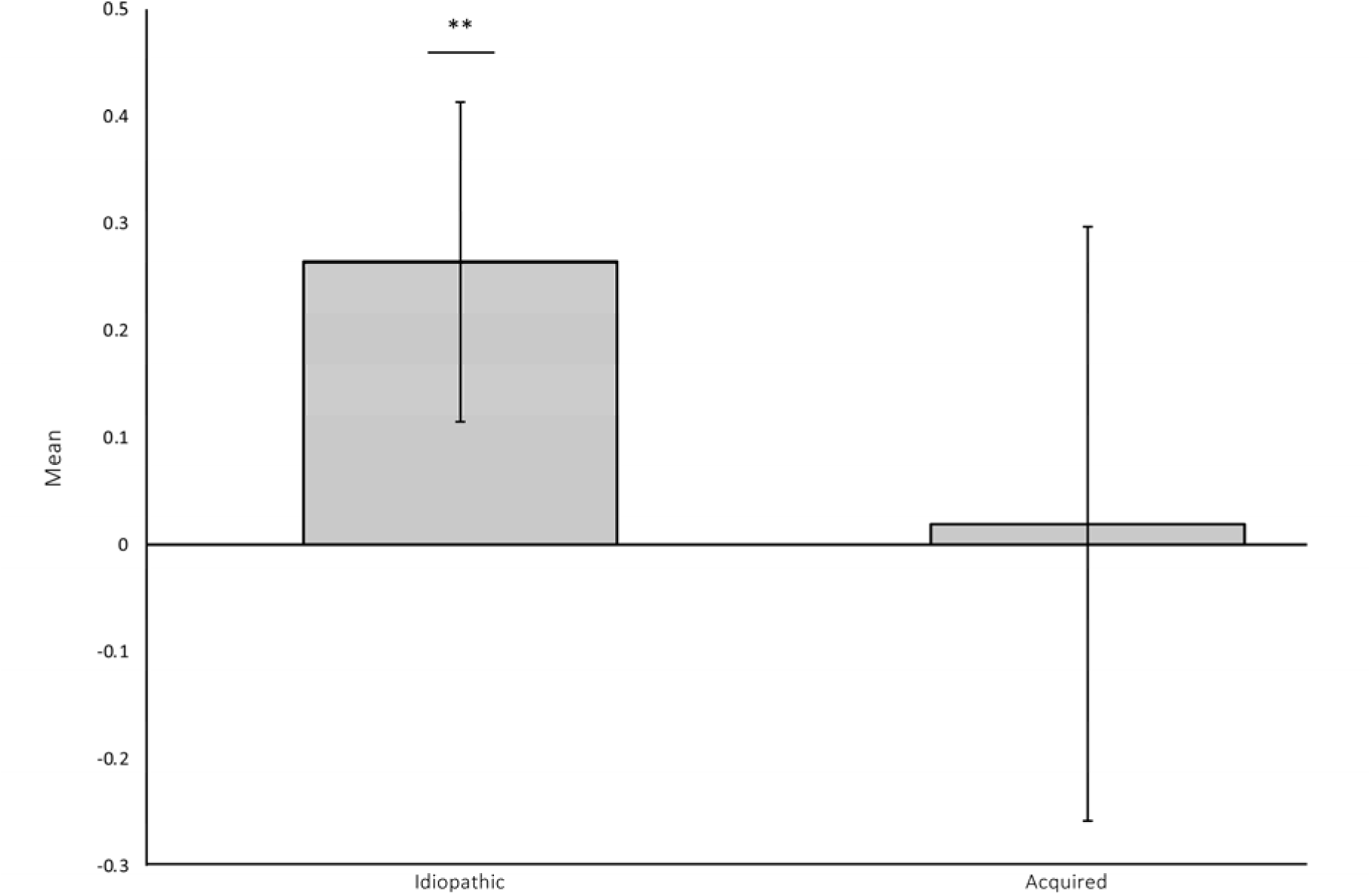
Meta-regression conducted on dystonia etiology. No significant differences were found between idiopathic and acquired dystonia, however, idiopathic dystonia effect sizes were significant. ** denotes significance and the *p* < .05level.

Effect sizes for studies which utilised motor training concurrently with NIBS were significantly larger than studies which applied NIBS alone, *Q*(1) = 4.43, *p* = .04. Overall mean effect sizes for both groups were significant, NIBS and motor training *g* = 0.55, *p* = .001 and NIBS alone *g* = 0.15, *p* = 0.03 (Figure 5).

**Figure 5.**
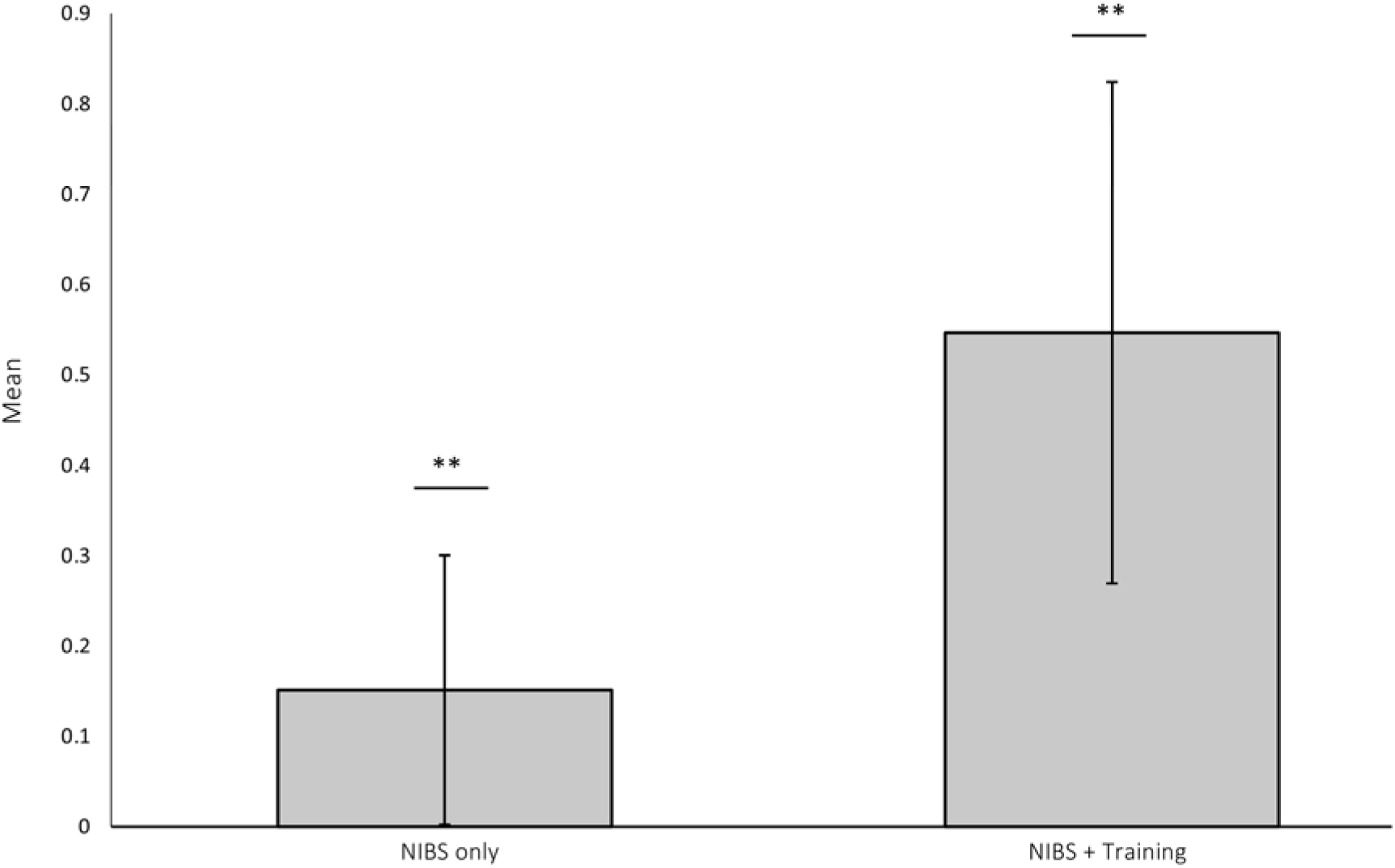
Meta-regression conducted on NIBS with and without concurrent motor training. No significant difference was found between NIBS and training and NIBS only, however, individual effects were significant. ** denotes significance and the *p* < .05 level.

Meta-regressions on mean age and gender ratio of participants were not significant: mean age *b* = 0.002, SE = 0.005, *p* = 0.66 and gender ratio *b* = -0.03, SE = 0.03, *p* = 0.28 (supplementary 11 and 12). Additionally, these moderators showed no significant effect when the outlier^64^ study was included (supplementary file 13 and 14).

### Evaluation of Bias

Methodological quality of studies, as assessed by the RoB2, is presented in Figure 6. An overall judgement of high risk of bias was given where studies had a high risk of bias in at least one domain. Three studies were considered to be at high risk of bias. Borich et al.^53^ was considered to be at high risk of bias due to missing outcome data. Bradnam et al.^55^ and Rossett-Llobet et al.^33^ indicated that participant allocation to sham or active NIBS group was not concealed, and thus were judged at high risk of bias for the domain of random sequence generation. Most studies were judged to be at an unclear risk of bias in the domain of random sequence generation due to a lack of reporting how participants were randomized, and whether the allocation sequence was concealed. Furthermore, the domain of selective outcome reporting was judged to be at an unclear risk of bias for most studies, due to insufficient information available to permit a judgement of low risk (e.g., trial protocols)^40^. Overall, the literature was characterized by an unclear risk of bias.

**Figure 6.**
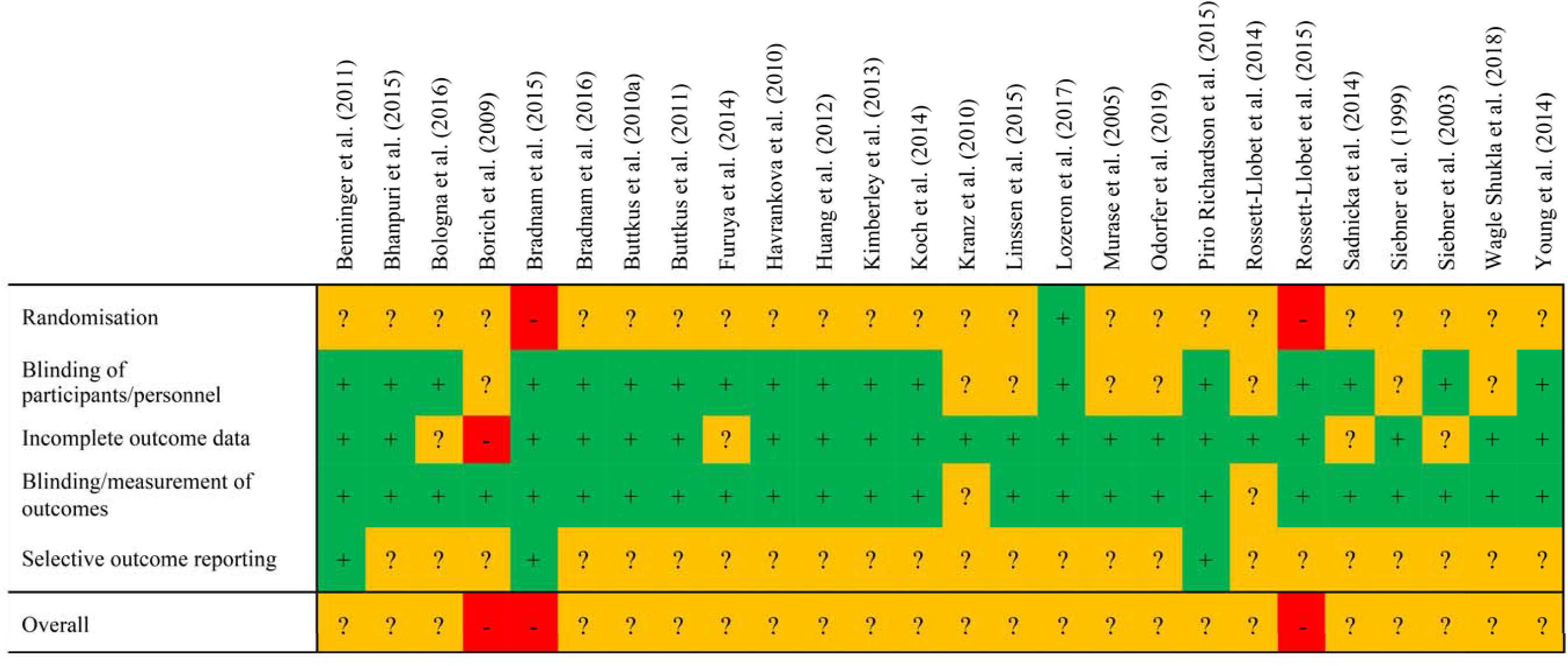
Risk of bias assessment for individual studies. Green boxes (+) = low risk of bias; orange boxes (?) = unclear risk of bias; red boxes (-) = high risk of bias.

The funnel plot analysis revealed two studies outside the boundaries of the funnel^32, 44^ (supplementary 15). Egger’s test trended towards significance (*t*(26) = 1.95*, p* = 0.06). Whilst this is indicative of symmetry within the funnel plot, suggesting that publication bias may not have affected this meta-analysis, results should be interpreted with caution given that an outlier study^64^ was not included in this analysis.

## Discussion

The primary aim of this systematic review and meta-analysis was to evaluate the efficacy of NIBS on dystonia symptoms. Overall meta-analysis of 27 studies demonstrated a small, yet significant effect for NIBS decreasing symptoms of dystonia. Further meta-analyses were then conducted separating studies by the different types of NIBS, dystonias, brain regions stimulated, and outcome measures. These analyses showed significantly reduced dystonia symptoms for 0.2Hz rTMS and cathodal tDCS, blepharospasm and task-specific FHD (including writer’s and musician’s dystonias individually),and the ACC, M1 and dPM. Lastly, meta-regression analyses suggested that 10 sessions of active stimulation, or NIBS applied concurrently with motor training had a significant effect on study effect size.

### Brain region stimulated and type of non-invasive brain stimulation

Studies stimulating the M1, dPM and ACC demonstrated significantly reduced dystonia symptoms. However, the ACC effect was only contributed to by one study, therefore this result should be interpreted with caution. Further, two inhibitory protocols were found increase the effect of NIBS – specifically, 0.2Hz rTMS and cathodal tDCS. That stimulation of the M1 and dPM, and use of inhibitory NIBS protocols significantly predicted an effect of NIBS on dystonia symptoms is in line with prior research, demonstrating increased excitability in sensorimotor areas including the motor, premotor and somatosensory cortices^15–17^. This can be seen through the excessive contraction of both agonist and antagonist muscles in dystonia, leading to unwanted muscle spasms and motor overflow^80^.Thus, the application of inhibitory NIBS protocols to these cortical areas may downregulate cortical and network activity, leading to a reduction in symptoms.

### Type of dystonia

When separating meta-analysis by type of dystonia, NIBS significantly reduced symptoms in blepharospasm and task-specific FHD, inclusive of musician’s dystonia and writer’s cramp. However, the effect for blepharospasm should be interpreted with caution, as only one study was included in this analysis^78^. While task-specific FHDs significantly benefitted from the application of NIBS, hand dystonia did not reach significance. Hand dystonia NIBS targets were spread over several brain regions, including the cerebellum and sensorimotor areas. Further, both inhibitory and excitatory NIBS protocols were used, with cTBS, anodal and cathodal tDCS, and 1Hz rTMS all trialed. The variability in protocol and targets in hand dystonia, along with the lack of contributing studies, is likely to have contributed to the non-significant finding. Conversely, task-specific FHD studies mainly targeted the M1 and dPM, with the most common NIBS protocol cathodal tDCS (or anodal and cathodal protocols combined in the same study) to the M1. Future trials in task-specific FHD should consider utilizing inhibitory protocols targeting the M1 and dPM, in order to maximize the therapeutic effects of NIBS in this cohort.

### Number of non-invasive brain stimulation sessions

Studies ranged from a single session of NIBS to several sessions over multiple weeks. 16 of the 20 studies included in the qualitative review applied multiple sessions of stimulation, all reporting a reduction in dystonia symptoms upon competition of the NIBS sessions – however, statistical significance for many studies was not reported. Meta-regression analysis demonstrated that 10 sessions of active stimulation was more effective for improving dystonia symptoms than one or five sessions of stimulation. The finding of 10 sessions of active stimulation having a larger mean effect than one session is consistent with previous research that suggests consecutive sessions of NIBS, such as rTMS, are more effective in inducing longer-lasting plastic changes within cortical regions such as the M1^81^. It is also consistent with clinical protocols for NIBS treatments in neuropsychiatric disorders where rTMS is applied over a number of sessions, for example, depression (30 sessions over 4-6 weeks)^82^ and obsessive-compulsive disorder (29 sessions)^83^. Nonetheless, optimal parameters for both NIBS protocols and session quantity and timing for dystonia are yet to be established. Future clinical trials should include at least 10 sessions of NIBS to increase therapeutic efficacy, and further examine cumulative effects of NIBS paradigms within dystonia patients.

### Concurrent non-invasive brain stimulation and motor training

There was a significant difference in effect sizes between studies which implemented concurrent NIBS and motor training and those where only NIBS was applied, with studies which applied concurrent NIBS and motor training having a larger overall effect on dystonia symptoms. All studies included in the meta-regression which implemented concurrent NIBS and motor training did so in musician’s dystonia patients, using tDCS to the M1. Studies utilised motor training programs such as sensory-motor retuning^32, 33^, a type of therapy commonly used in musician’s dystonia that facilitates proprioceptive changes in the affected limb, and helps to modify abnormal cortical organization of sensory areas^84^. Research in stroke patients indicates that utilizing tDCS over the sensorimotor areas in conjunction with motor training can improve motor function and produce functional changes in sensorimotor areas beyond that of training alone^85–87^. The use of tDCS may assist with improvement of motor functioning by modulating cortical excitability and increasing plasticity within the targeted cortical area, allowing for optimal conditions in which to consolidate the effects of motor training or therapy^88^. Thus, future research should further examine the promising therapeutic effects of combined tDCS and motor training programs, such as sensory-motor retuning, in other types of dystonia beyond musician’s dystonia.

### Idiopathic versus acquired dystonia

Meta-regression demonstrated that, although there was no significant difference between idiopathic and acquired dystonia study effect sizes, idiopathic dystonia studies had a significant mean effect. Of the studies that utilised acquired dystonia patients in the overall meta-analysis, two studies recruited participants with cerebral palsy^44, 45^ and one with Wilson’s disease^43^. Given that the basal ganglia are thought to be involved in dystonia as part of the sensorimotor network, the atrophy or lesioning of this brain region, as is often seen in cerebral palsy and Wilson’s disease patients, may result in different NIBS outcomes for those with acquired dystonia in comparison to those with idiopathic dystonia. Previous research in idiopathic writer’s cramp patients has demonstrated reduced functional connectivity in comparison to healthy controls, in areas such as the bilateral thalamus, putamen and globus pallidus, and left dPM^89^. However, a single session of rTMS induced a significant increase in connectivity in basal ganglia regions, specifically the bilateral thalamus and putamen^89^. This suggests that although NIBS is applied to the cortex, effects extend to the basal ganglia and other subcortical structures, highlighting the need for an unaltered pathway between basal ganglia and stimulated cortex in dystonia patients to optimize NIBS outcomes^45^.

## Limitations

A limitation of this meta-analysis was only reviewing dystonia outcomes at the first time-point of assessment after the NIBS intervention. Several studies examined the effects of the NIBS at multiple timepoints (e.g., mid-intervention or four weeks post-intervention), and thus only estimating the effect of NIBS at the immediate end point of the intervention may have led to an overestimation of the true intervention effect, and may not accurately inform how effective the use of NIBS on symptoms of dystonia is long-term.

A moderate level of between-study heterogeneity was found in this meta-analysis. Whilst secondary analyses were conducted to find moderators of the effect, other methodological differences between studies may have contributed to the significant level of heterogeneity – for example, the number of pulses applied in rTMS protocols. The overall methodological quality of the evidence was mixed, with Figure 6 demonstrating the uncertainty in whether randomization and selective outcome reporting influenced individual study results, and thus overall effect size. Notably, the inability to judge the domain of selective outcome reporting as low risk may suggest that the study-level effect sizes were, to a degree, overestimated. Although Egger’s test was non-significant, suggesting that the research field may not suffer from publication bias, meta-analysis results should be considered bearing in mind the standard of reporting.

## Conclusions

The present systematic review and meta-analysis found a small effect size in favor of NIBS reducing symptoms of dystonia. The use of ‘inhibitory’ NIBS protocols (i.e., 0.2Hz rTMS and cathodal tDCS), stimulation of the M1 and dPM, protocols employing a greater number of sessions, and concurrent motor training protocols demonstrated the highest treatment effects for NIBS. Future research should apply 10 sessions or more of NIBS and further investigate the use of motor training concurrently with NIBS, to yield the high-quality evidence needed to translate this promising therapeutic technique to clinical use.

## Data Availability

All data in the present study are available upon reasonable request to the authors.

## Author roles

Jordan Morrison-Ham 1abc, 2ab, 3ab.

Gillian M. Clark 2abc, 3b.

Elizabeth G. Ellis 1c, 3b.

Andris Cerins 1c, 3b.

Juho Joutsa 1ab, 3c.

Peter G. Enticott 1ab, 3c.

Daniel T. Corp 1abc, 2abc, 3b.

1. **Research Project.** a. Conception, b. organization, c. execution.
2. **Statistical Analysis.** a. design, b. execution, c. review/critique.
3. **Manuscript.** a. writing of the first draft, b. review/critique.

## Acronyms

ACC: anterior cingulate cortex
CD: cervical dystonia
dPM: dorsal premotor cortex
FHD: focal hand dystonia
M1: primary motor cortex
NIBS: non-invasive brain stimulation
rTMS: repetitive transcranial magnetic stimulation
S1: primary somatosensory cortex
TBS: theta-burst stimulation
tDCS: transcranial direct current stimulation
TMS: transcranial magnetic stimulation.

## Notes

**Funding/Conflicts of interest:** JMH, EGE & AC are funded by an Australian Government Research Training Program Scholarship. JJ is funded by the Instrumentarium Research Foundation, the Finnish Foundation for Alcohol Studies, University of Turku (private donation, Sigrid Juselius Foundation) and Turku University Hospital (ERVA funds), and has received a lecturer honorarium from Lundbeck. PGE is funded by a Future Fellowship from the Australian Research Council (FT160100077).

### Competing Interest Statement

The authors have declared no competing interest.

### Funding Statement

JMH, EGE & AC are funded by an Australian Government Research Training Program Scholarship. JJ is funded by the Instrumentarium Research Foundation, the Finnish Foundation for Alcohol Studies, University of Turku (private donation, Sigrid Juselius Foundation) and Turku University Hospital (ERVA funds), and has received a lecturer honorarium from Lundbeck. PGE is funded by a Future Fellowship from the Australian Research Council (FT160100077).

### Author Declarations

The data used in the production of this manuscript were obtained from existing studies and used no human subjects.

