## Supplementary 15 - funnel plot analysis for "Effects of non-invasive brain stimulation in dystonia: a systematic review and meta-analysis" (from another version of this preprint: version 1, 2021-11-03)

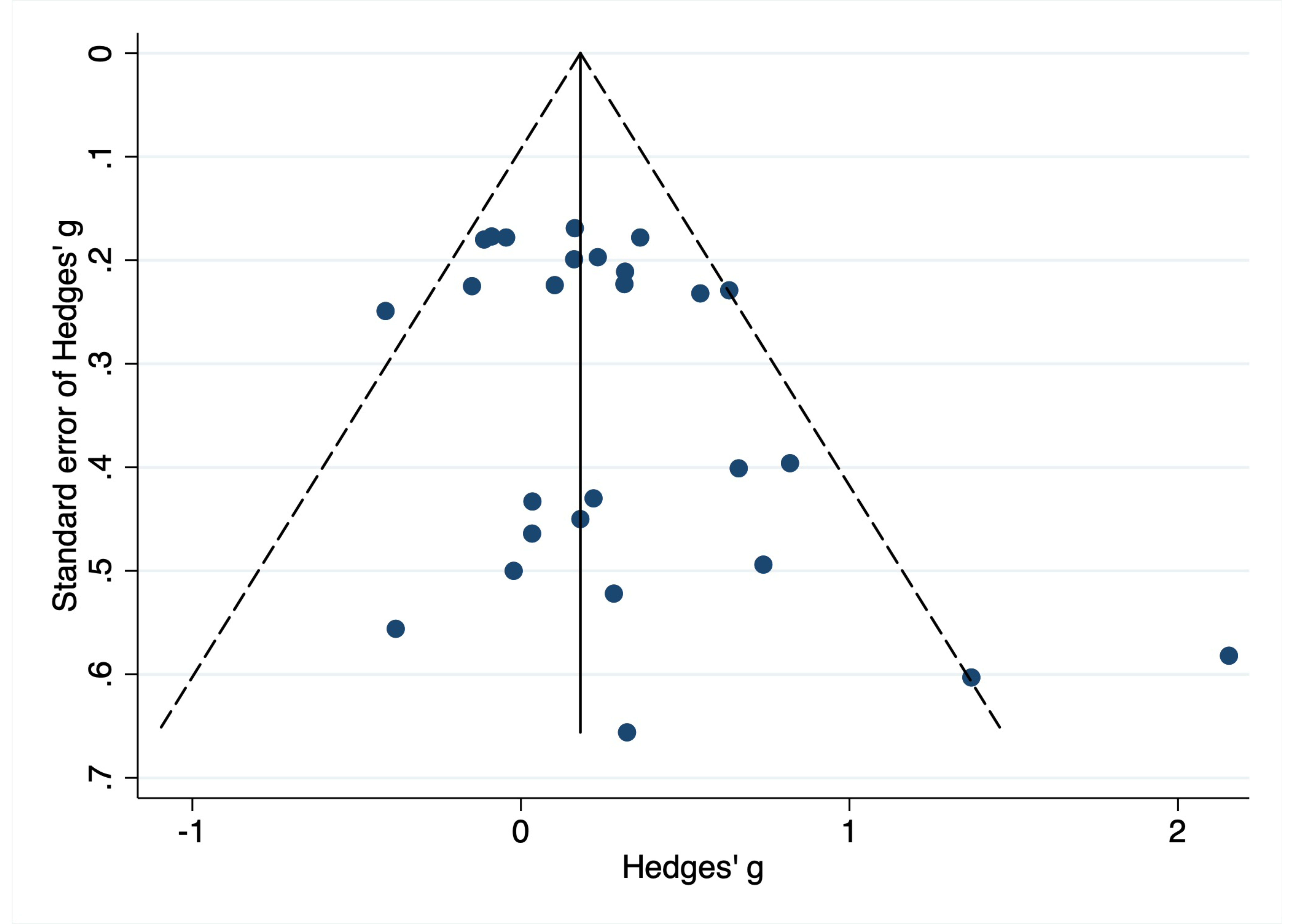

*Note.* Funnel plot analysis examining publication bias across studies. Two studies were outside the boundaries of the funnel^32, 44^.
