## Supplementary 13 - meta-regression mean age (with outlier) for "Effects of non-invasive brain stimulation in dystonia: a systematic review and meta-analysis" (from another version of this preprint: version 1, 2021-11-03)

**Supplementary 13 – Meta-regression on mean age (outlier included)**
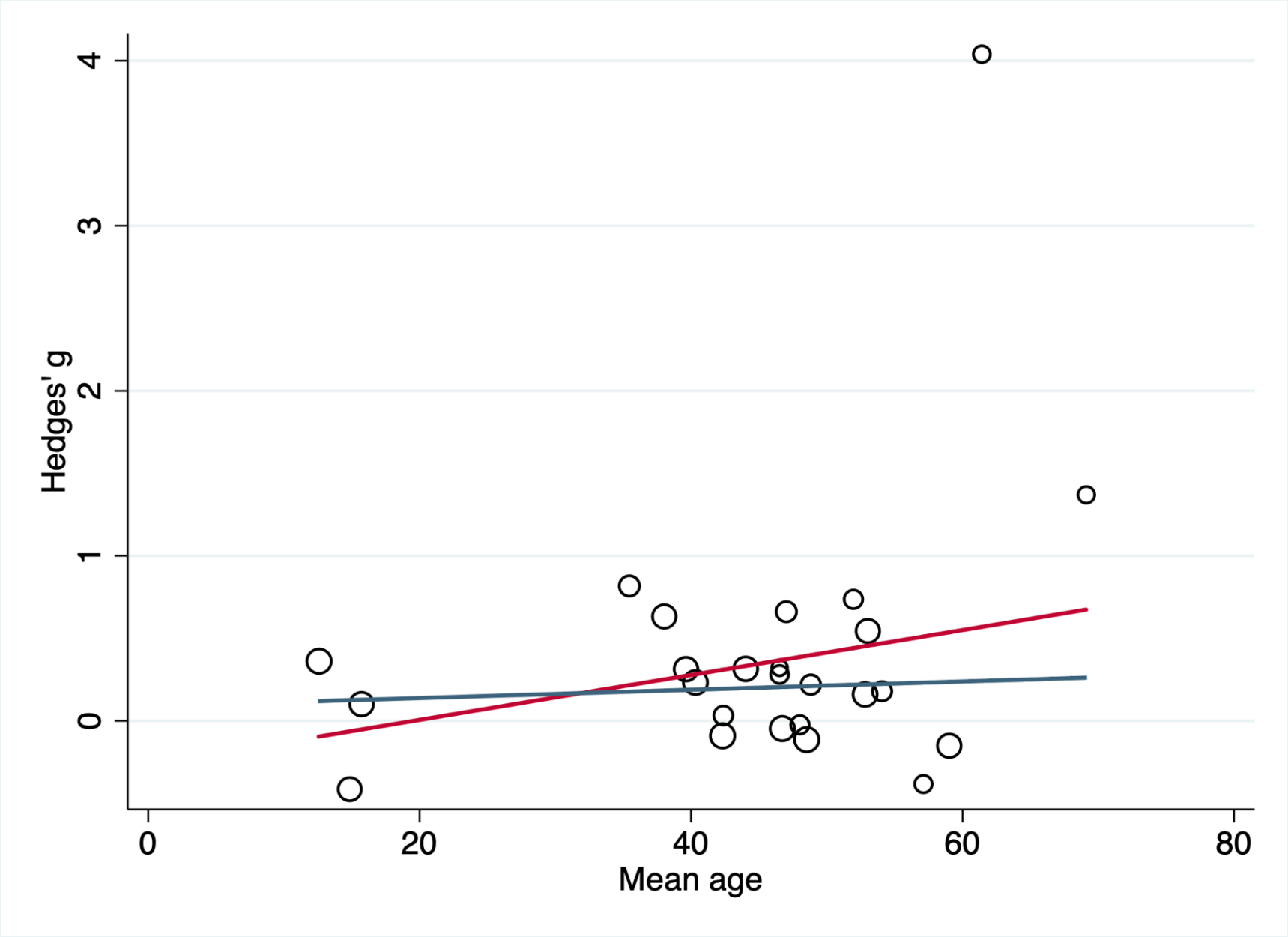

*Note.* The size of the circles reflects the weighting given to each study included within the meta-regression. The blue line represents the line of best fit with Kranz et al. (2010) removed, and the red where all studies are included.
