## Supplementary 12 - meta-regression gender for "Effects of non-invasive brain stimulation in dystonia: a systematic review and meta-analysis" (from another version of this preprint: version 1, 2021-11-03)

**Supplementary 12 – Meta-regression on gender ratio**

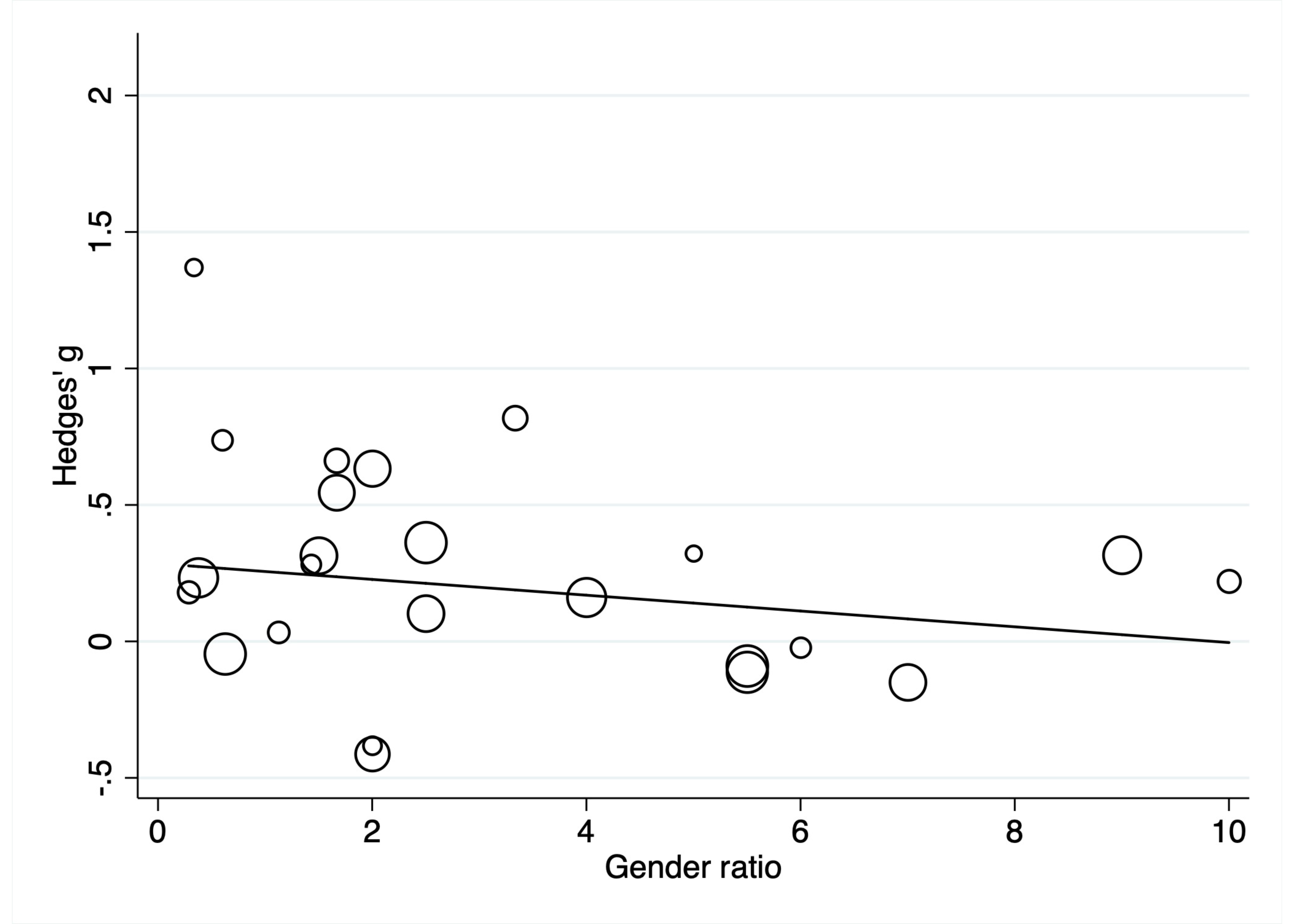

*Note.* Meta-regression of gender ratio on Hedges’ *g* effect sizes. The x-axis represents gender ratio in studies, indicating the number of male participants per female participant in the study. The size of the circles reflects the weighting given to each study included within the meta-regression.
