## Supplementary 11 - meta-regression mean age for "Effects of non-invasive brain stimulation in dystonia: a systematic review and meta-analysis" (from another version of this preprint: version 1, 2021-11-03)

**Supplementary 11 – Meta-regression on mean age**

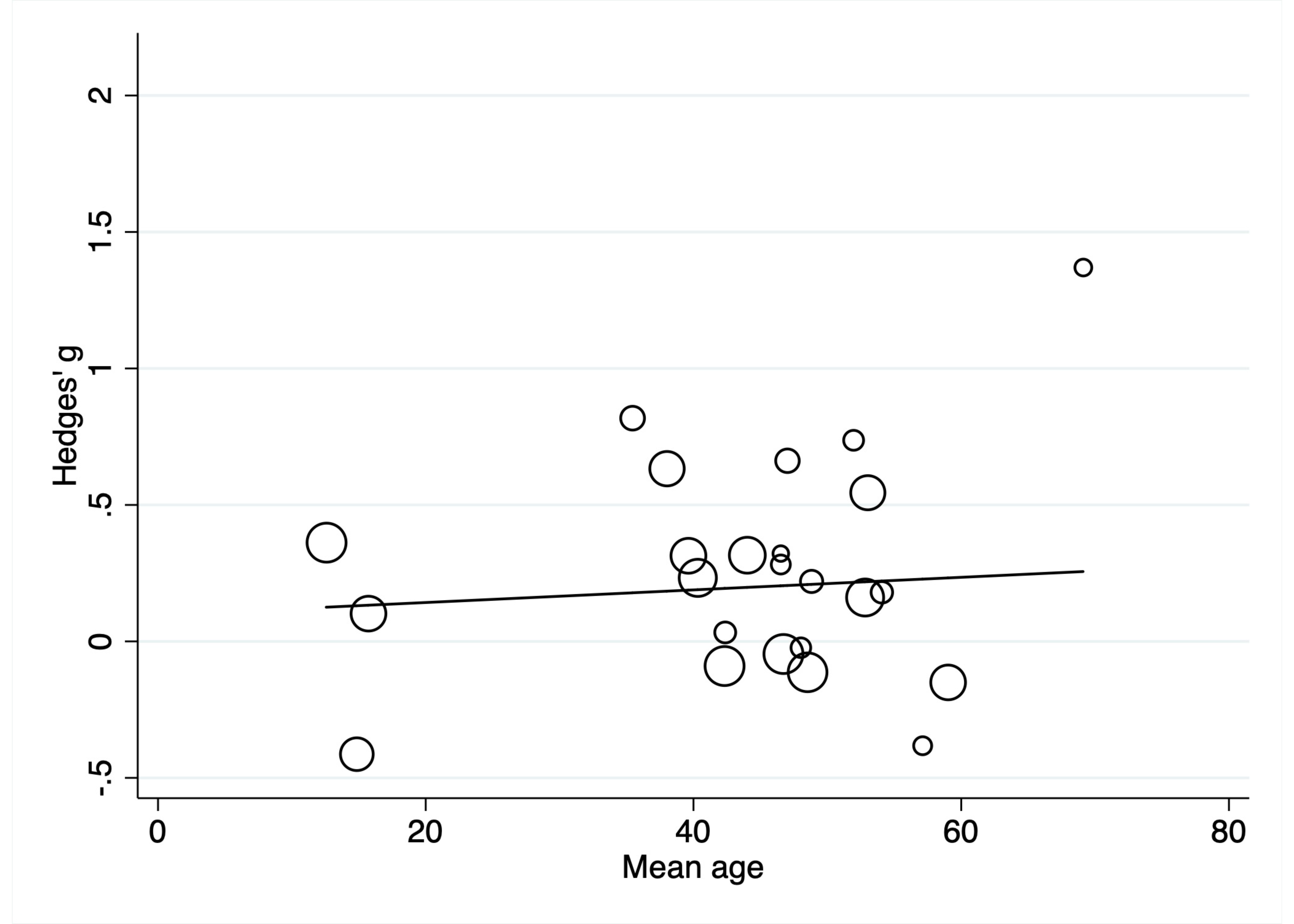

*Note.* Meta-regression of mean age on Hedges’ *g* effect sizes. The x-axis represents mean age in years. The size of the circles reflects the weighting given to each study included within the meta-regression.
