## Supplementary figures and images for "Effects of non-invasive brain stimulation in dystonia: a systematic review and meta-analysis" (from another version of this preprint: version 1, 2021-11-03)

### Supplementary 3 - leave-one-out analysis

**Supplementary 3 – Leave-one-out meta-analysis**
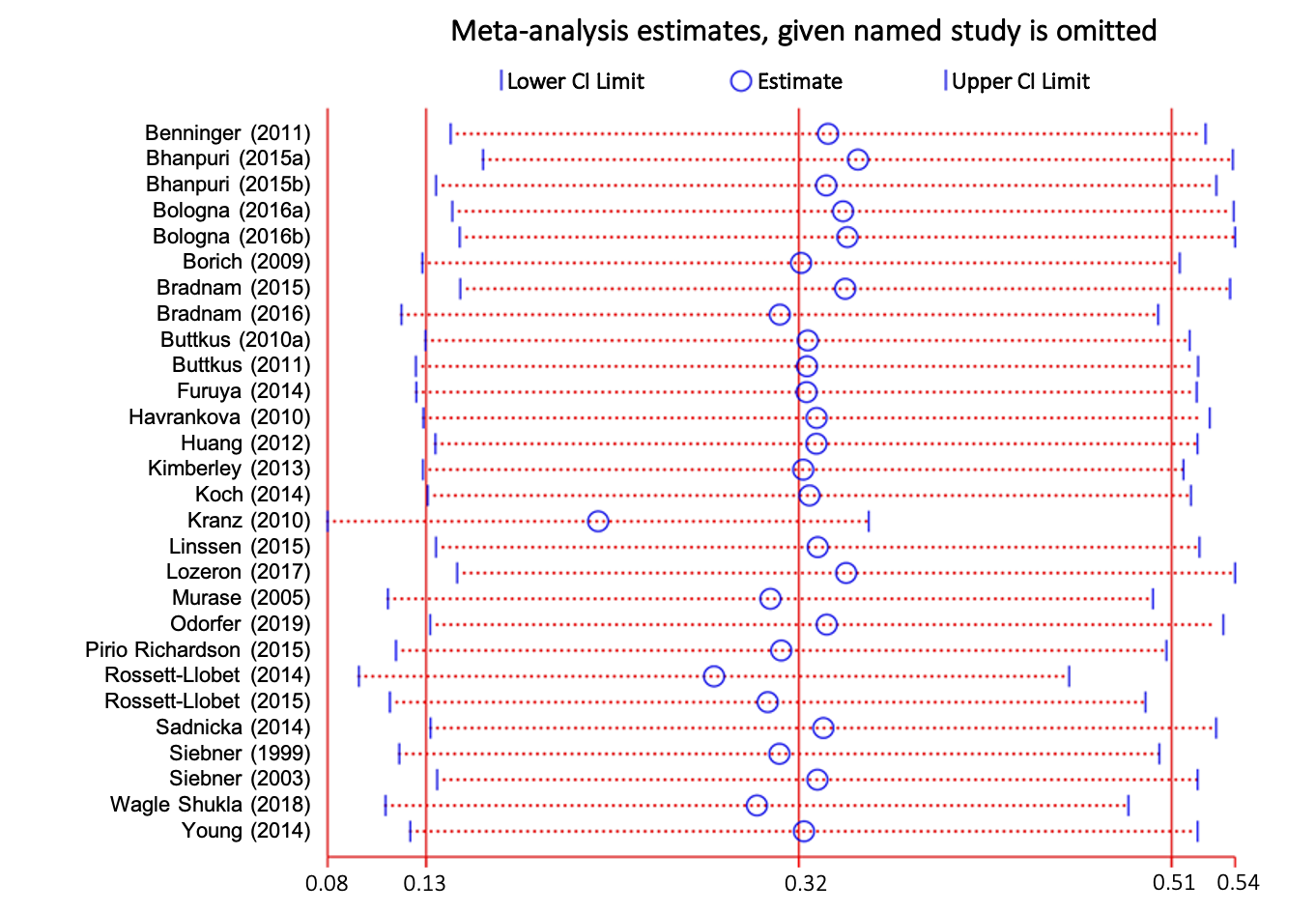

### Supplementary 4 - meta-analysis

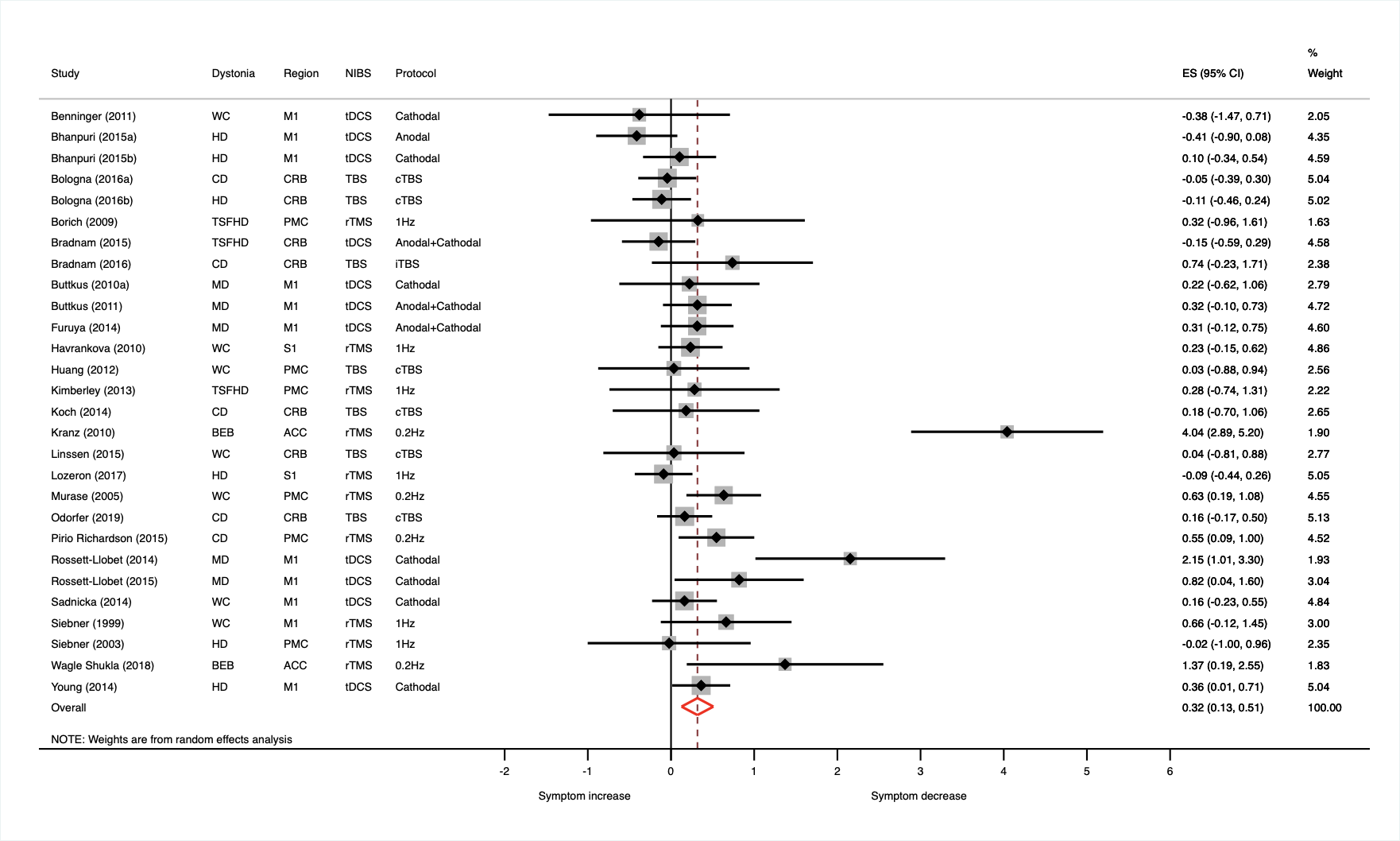
**Supplementary 4 – Meta-analysis with outlier included**

### Supplementary 5 - forest plot NIBS

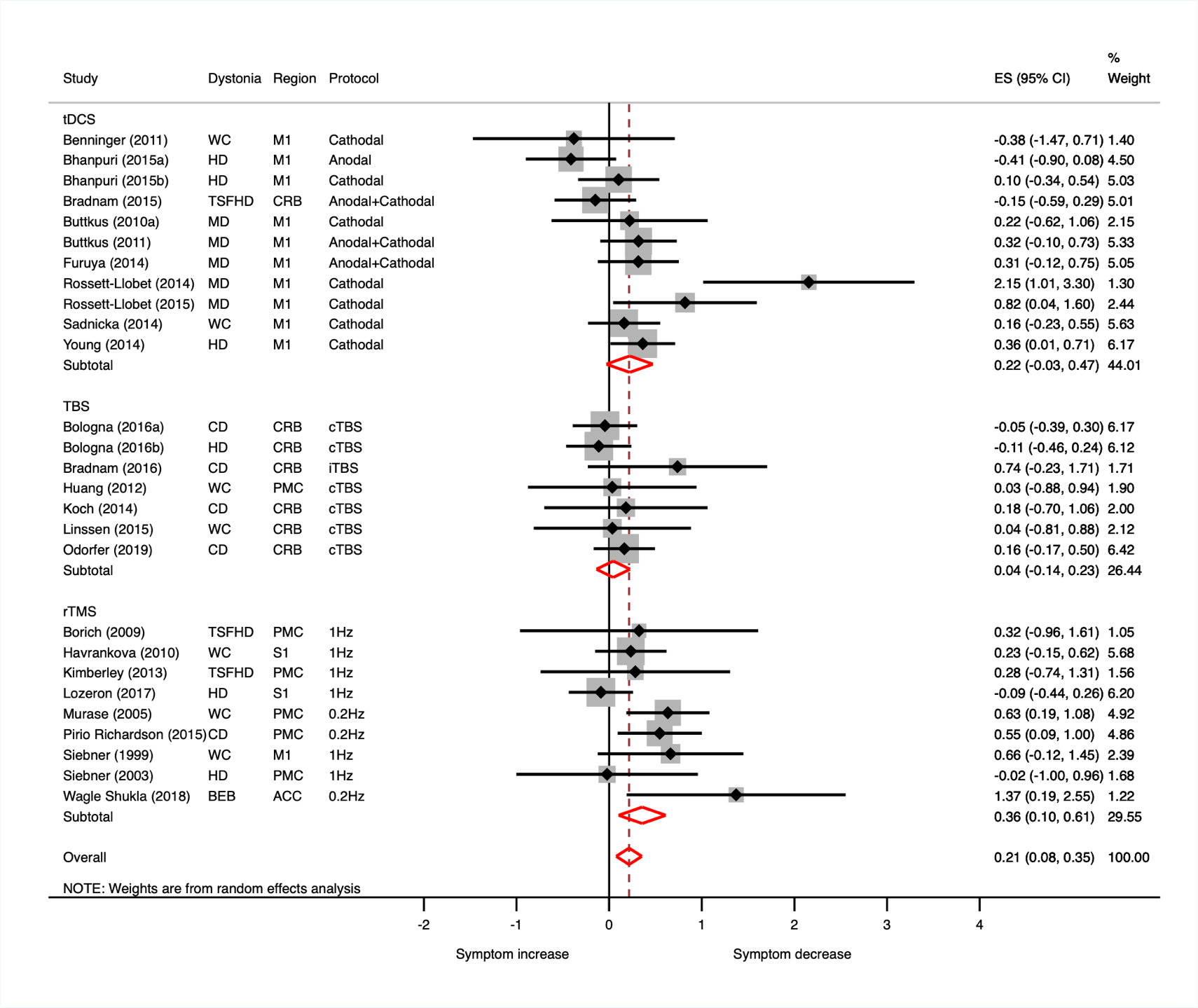
**Supplementary 5 – Forest plot separating studies by type of NIBS**

### Supplementary 6 - forest plot type of dystonia

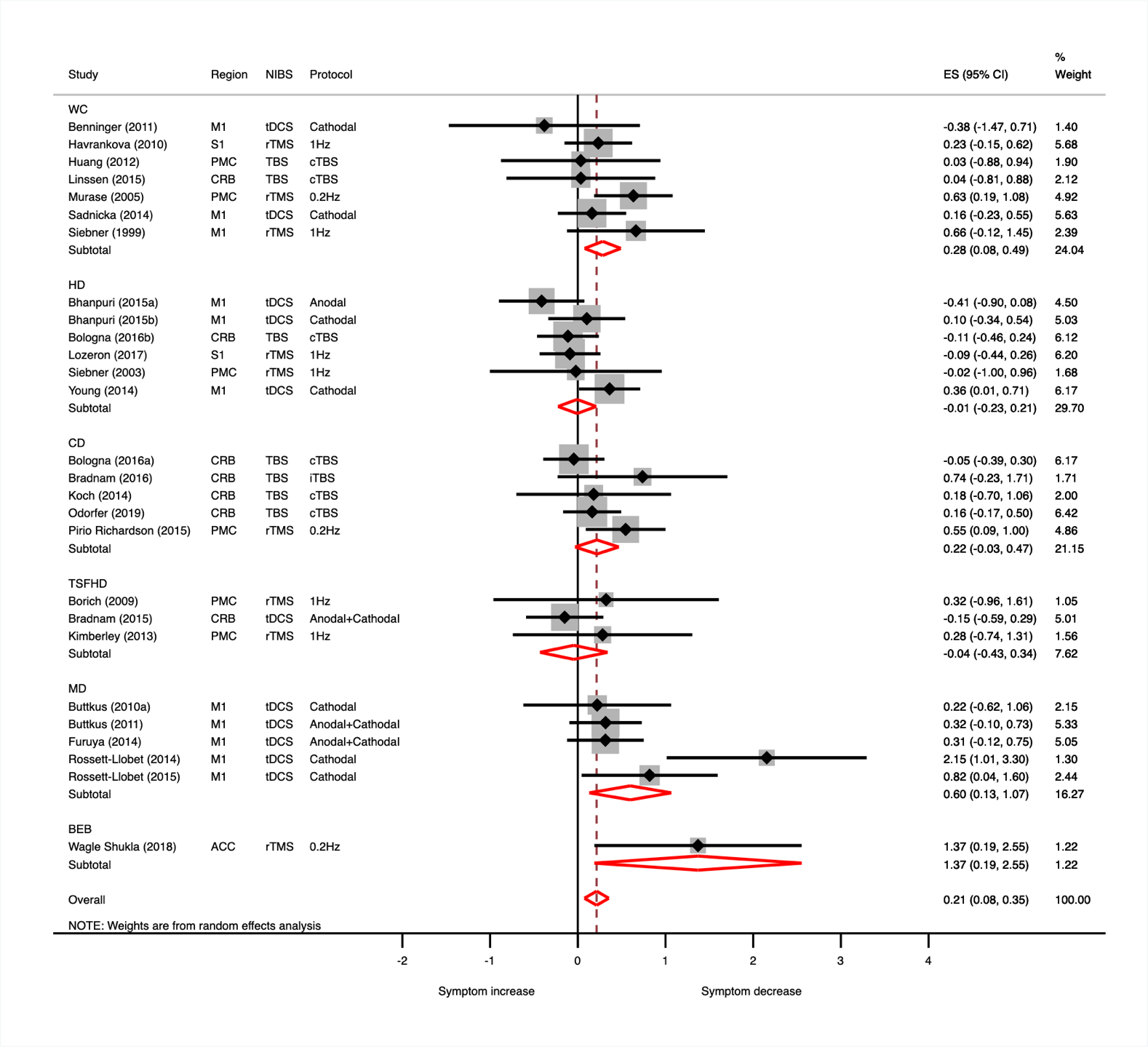
**Supplementary 6 – Forest plot separating studies by type of dystonia**

### Supplementary 7 - forest plot brain region

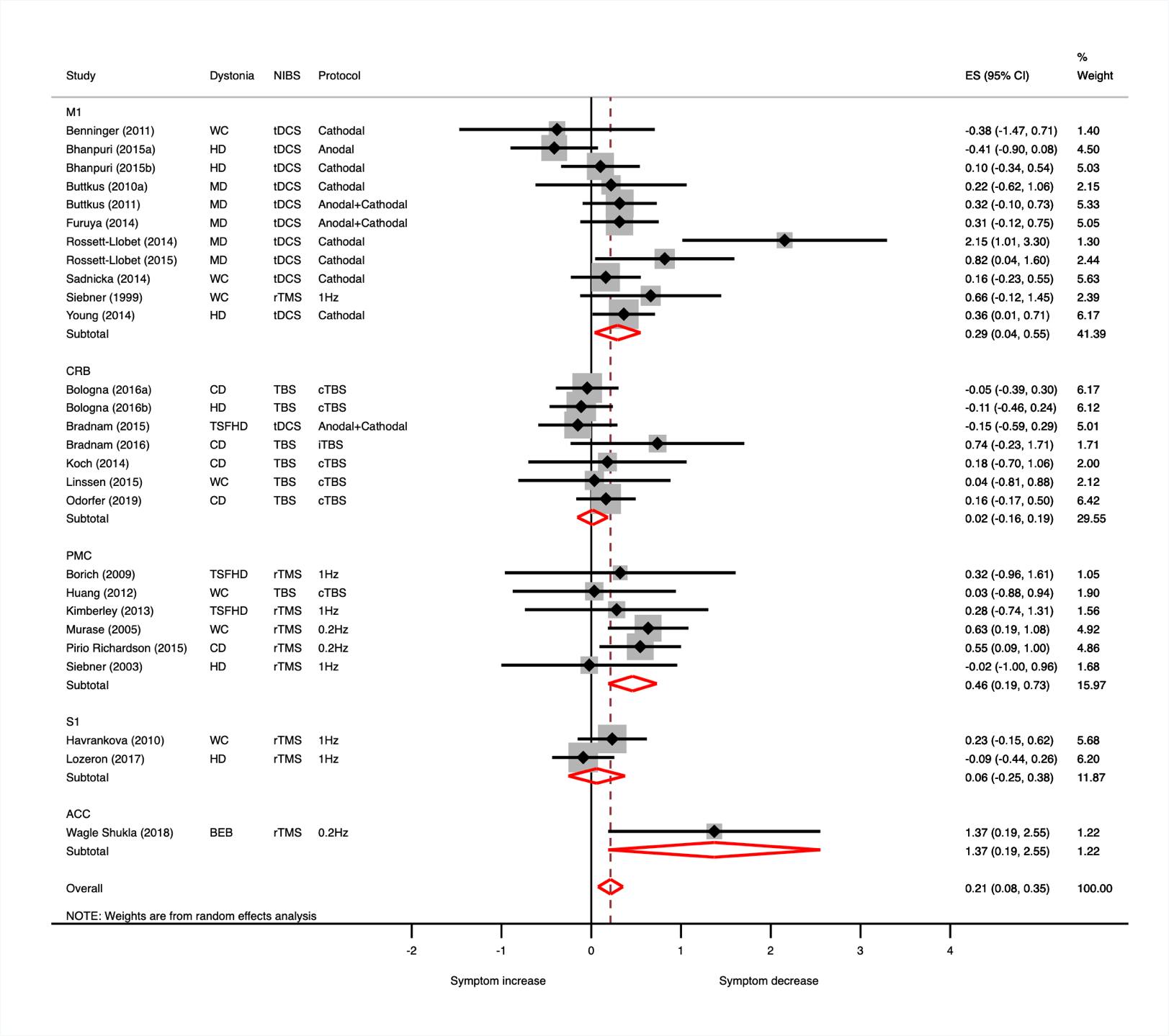
**Supplementary 7 – Forest plot separating studies by brain region**

### Supplementary 8 - forest plot validates scales

**Supplementary 8 – Forest plot of studies using validated scale outcomes**
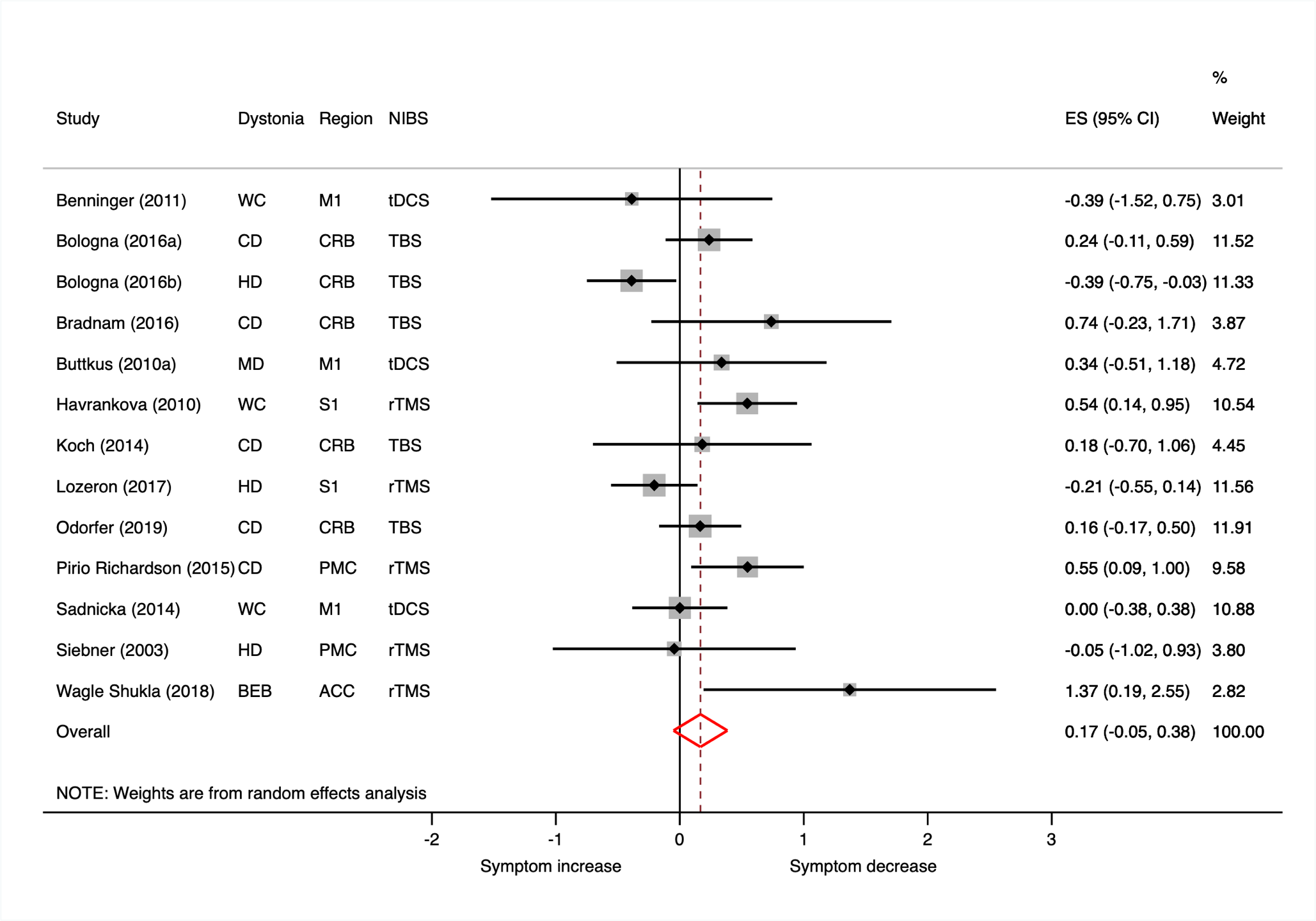

### Supplementary 9 - forest plot unvalidated scales

**Supplementary 9 – Forest plot of studies using unvalidated scale outcomes**

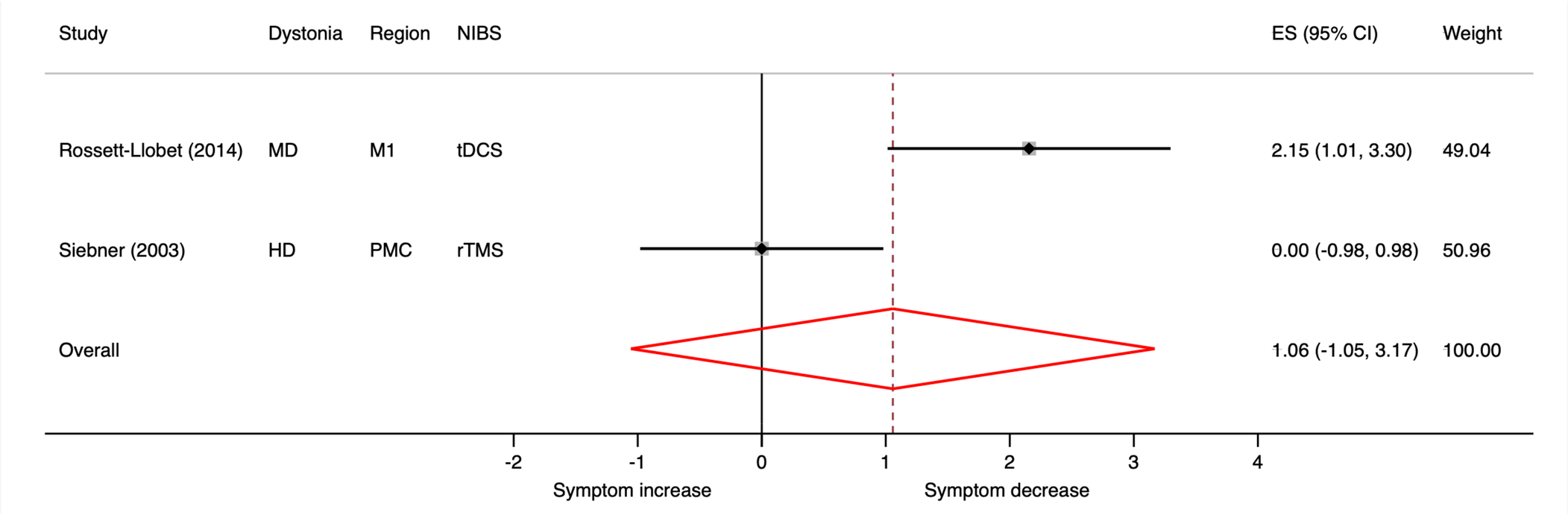

### Supplementary 10 - forest plot task-based measures

**Supplementary 10– Forest plot of studies using task-based outcomes**
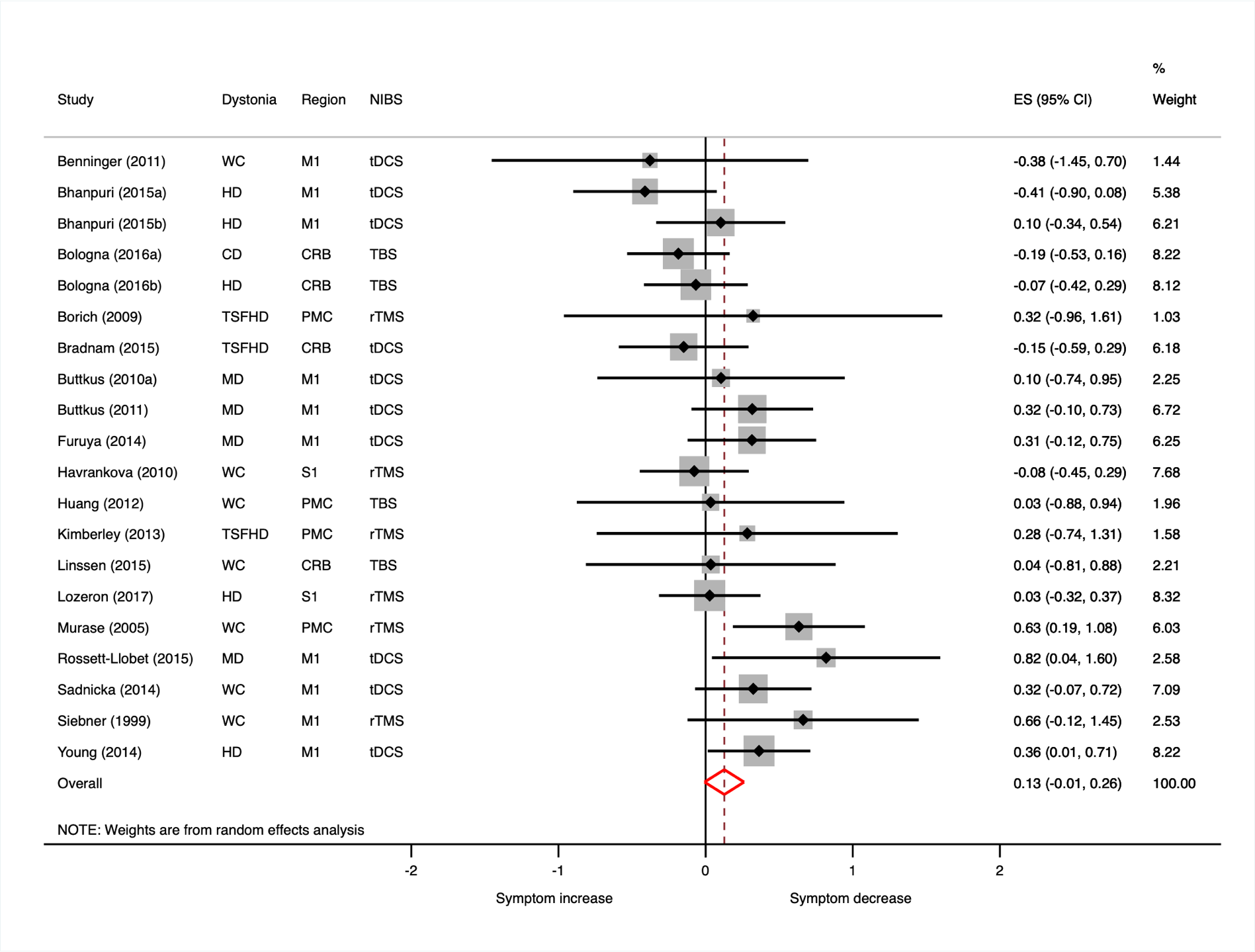
