## Supplementary 2 - effect size calculations for "Effects of non-invasive brain stimulation in dystonia: a systematic review and meta-analysis" (from another version of this preprint: version 1, 2021-11-03)

*Parallel Trials*

For parallel groups trials the SMD was calculated using the CMA (version 3.3.070) ‘unmatched groups, pre and post data’ function, with the option to standardise by change standard deviation (SD) selected. The formula is as follows:

$$SMD= \frac{\bar{X}_{changeE}- \bar{X}_{changeC}}{{SD}_{pooled}} (1)$$

Where:

$${SD}_{pooled}= \frac{\sqrt{(\left( n_{E}-1 \right)*{({SD}_{changeE}}^{2}))+(\left( n_{C}-1 \right)*({{SD}_{changeC}}^{2})})}{n_{E}+ n_{C}-2} (2)$$

$${SD}_{change}= \sqrt{{{({SD}_{pre}}^{2})+({SD}_{post}}^{2})-(2*r*{SD}_{pre}* {SD}_{post})} (3)$$

Where E represents the experimental or NIBS group, C represents the sham stimulation group, *n* represents the number of participants within the group, and *r* represents the correlation coefficient across groups (i.e., how similar the baseline and post-intervention measurements were across groups of participants). As the correlation coefficient was not reported in any studies, a weighted average correlation of 0.77 was obtained using individual patient data from five studies^29, 31-33, 53^, and utilised for the calculation of both parallel and crossover group trial effect sizes. For studies that reported an *F* value, this was imputed in CMA and a resulting SMD was obtained with the formula:

$$SMD= \sqrt{\frac{F*(n_{E}+ n_{C})}{n_{E}* n_{C}}} (4)$$

The standard error (SE) for the SMD was obtained by the formula:

$${SE}_{SMD}= \sqrt{\frac{\left( \frac{1}{n_{E}} \right)+\left( \frac{1}{n_{C}} \right)+({{SD}_{pooled}}^{2})}{2*(n_{E}+n_{C)}}} (5)$$

SMD and SE values were then multiplied by the correction factor, *J*, to obtain a Hedges’ *g* and corresponding SE:

$$J=1-\frac{3}{\left( 4*df \right)-1} (6)$$

Where *df* for parallel groups is obtained by the formula:

$$df= n_{E}+ n_{C}-2 (7)$$

*Crossover Trials*

For crossover group trials the SMD was calculated using the CMA one-group (pre-post) and matched groups function, using formula 1. ${SD}_{pooled}$ represents the SD of the change from baseline scores between groups, and is calculated for crossover trials using the formula:

$${SD}_{pooled}=\sqrt{\frac{{SD}_{diff}}{\sqrt{2*\left( 1-r \right)}}} (8)$$

Where:

$${SD}_{diff}=\sqrt{{{(SD}_{changeC}}^{2})+\left( {{SD}_{changeE}}^{2} \right)-(2*r*{SD}_{changeC}*{SD}_{changeE})} (9)$$

For several studies^29, 30, 52, 55, 58, 64, 68^ the mean change score and SD of the change score was not given, and thus was calculated from the difference between pre- and post-stimulation (or sham) mean scores^28^. The SD of the change score was calculated for each group using the formula^28^:

$${SD}_{change}= \sqrt{{{(SD}_{pre}}^{2})+\left( {{SD}_{post}}^{2} \right)-(2*r*{SD}_{pre}*{SD}_{post})} (10)$$

The SE for the SMD for crossover group trials was calculated using the formula^28^:

$$S{MD}_{SE}= \sqrt{\left( \frac{1}{N} \right)+\frac{{SMD}^{2}}{2N}} *\sqrt{2*(1-r)} (12)$$

SMD and SE scores were then multiplied by the correction factor *J* (6), to obtain a Hedge’s *g* and corresponding SE value. In crossover trials, the *df* value for the correction factor, *J*, is obtained by:

$$df=N-1 (13)$$

Where *N* is the total number of participants in the study.
