## Supplementary 1 - search syntax for "Effects of non-invasive brain stimulation in dystonia: a systematic review and meta-analysis" (from another version of this preprint: version 1, 2021-11-03)

**Embase**
S1: dystoni*:ab,ti OR torticollis:ab,ti OR blepharospasm:ab,ti OR meige*:ab,ti OR 'writ* cramp':ab,ti OR 'dystonia'/de OR 'torticollis'/de OR 'blepharospasm'/de OR 'meige disease’/de

S2: 'transcranial magnetic stimulation':ab,ti OR tms:ab,ti OR tbs:ab,ti OR 'theta burst stimulation':ab,ti OR 'theta-burst stimulation':ab,ti OR tacs:ab,ti OR 'transcranial alternating current stimulation':ab,ti OR tdcs:ab,ti OR 'transcranial direct current stimulation':ab,ti OR tes:ab,ti OR 'transcranial electrical stimulation':ab,ti OR nibs:ab,ti OR 'noninvasive brain stimulation':ab,ti OR 'non-invasive brain stimulation':ab,ti OR 'transcranial random noise stimulation':ab,ti OR trns:ab,ti OR 'transcranial magnetic stimulation'/de OR 'theta burst stimulation'/de OR 'noninvasive brain stimulation'/de OR 'transcranial electrical stimulation'/de OR 'transcranial direct current stimulation'/de OR 'transcranial alternating current stimulation'/de OR 'transcranial random noise stimulation'/de

S3: S1 AND S2
Results = 794

**MEDLINE Complete**
S1: TI (dystoni* OR torticollis OR blepharospasm OR meige* OR writ* cramp) OR AB (dystoni* OR torticollis OR blepharospasm OR meige* OR writ* cramp)

S2: TI (transcranial magnetic stimulation OR TMS OR TBS OR theta burst stimulation OR theta-burst stimulation OR tACS OR transcranial alternating current stimulation OR tDCS OR transcranial direct current stimulation OR TES OR transcranial electrical stimulation OR NIBS OR noninvasive brain stimulation OR non-invasive brain stimulation OR transcranial random noise stimulation OR trns) OR AB (transcranial magnetic stimulation OR TMS OR TBS OR theta burst stimulation OR theta-burst stimulation OR tACS OR transcranial alternating current stimulation OR tDCS OR transcranial direct current stimulation OR TES OR transcranial electrical stimulation OR NIBS OR noninvasive brain stimulation OR non-invasive brain stimulation OR transcranial random noise stimulation OR trns)

S3: S1 AND S2
Results = 330
